# A Data Driven Approach for Choosing a Wearable Sleep Tracker

**DOI:** 10.1101/2023.10.12.23296981

**Authors:** Ju Lynn Ong, Hosein Aghayan Golkashani, Shohreh Ghorbani, Kian F Wong, Nicholas IYN Chee, Adrian R Willoughby, Michael WL Chee

## Abstract

**Goal and Aims:** To evaluate the performance of 6 wearable devices across 4 device classes (research-grade EEG-based headband, research-grade actigraphy, high-end consumer tracker, low-cost consumer tracker) over 3 age-groups (young: 18-30y, middle-aged: 31-50y and older adults: 51-70y).

**Focus Technology:** Dreem 3 headband, Actigraph GT9X, Oura ring Gen3 running the latest sleep staging algorithm (OSSA 2.0), Fitbit Sense, Xiaomi Mi Band 7, Axtro Fit3.

**Reference Technology:** In-lab polysomnography (PSG) with consensus sleep scoring.

**Sample:** 60 participants (26 males) across 3 age groups (young: N=21, middle-aged: N=23 and older adults: N=16).

**Design:** Participants slept overnight in a sleep laboratory from their habitual sleep time to wake time, wearing 5 devices concurrently.

**Core Analytics:** Discrepancy and epoch-by-epoch analyses for sleep/wake (2-stage) and sleep-stage (4-stage; wake/light/deep/REM) classification (devices vs. PSG). Mixed model ANOVAs for comparisons of biases across devices (within-subject), and age and sex (between-subjects).

**Core Outcomes:** The EEG-based Dreem headband outperformed the other wearables in terms of 2-stage (kappa = .76) and 4-stage (kappa = .76-.86) classification but was not tolerated by at least 25% of participants. This was followed by the high-end, validated consumer trackers: Oura (2-stage kappa = .64, 4-stage kappa = .55-.70) and Fitbit (2-stage kappa = .58, 4-stage kappa = .45-.60). Next was the accelerometry-based research-grade Actigraph which only provided 2-stage classification (kappa = .47), and finally the low-cost consumer trackers which had very low kappa values overall (2-stage kappa < .31, 4-stage kappa < .33).

**Important Additional Outcomes:** Proportional biases were driven by nights with poorer sleep (i.e., longer sleep onset latencies [SOL] and wake after sleep onset [WASO]). For those nights with sleep efficiency ≥85%, the large majority of sleep measure estimates from Dreem, Oura, Fitbit and Actigraph were within clinically acceptable limits of 30 mins. Biases for total sleep time [TST] and WASO were also largest in older participants who tended to have poorer sleep.

**Core Conclusion:** The Dreem band is recommended for highest accuracy sleep tracking, but it has price, comfort and ease of use trade-offs. The high-end consumer sleep trackers (Oura, Fitbit) balance classification accuracy with cost, comfort and ease of use and are recommended for large-scale population studies where sleep is mostly normal. The low-cost trackers, despite poor wake detection could have some utility for logging time in bed.

## INTRODUCTION

Increased awareness that sleep is a modifiable lifestyle factor for health and wellbeing has contributed to explosive growth in the sales of consumer wearable devices. Sales for digital fitness and wellbeing devices were valued at $58.11 billion worldwide in 2022 and are projected to grow at a compounded rate of ∼10% a year for at least 5 years.^1^ With many options to choose from, how does one decide on what to buy? Online polls conducted by sleep scientists show that both researchers and consumers value devices whose performance has been endorsed by experts.^2^ However, consumers and product reviewers often forgo scientific considerations for features, user experience or price.^3^ Also, interest in tracking daily physical activity (∼43% of wearable device users) outweighs interest in sleep tracking (∼19%) for users in the US,^4^ and physical activity improvements were more commonly sustained than diet or sleep.^5^ Finally, the US Food and Drug Administration and other medical regulators presently do not enforce minimal quality standards for consumer devices. Together, these market factors remove incentives for manufacturers to refine sleep assessment technology beyond that which is sufficient to maintain profitability. This contributes significantly to the heterogeneity in sleep quality assessment from different devices.

At the higher end of the market, well-conducted performance evaluation studies have shown that some consumer sleep trackers (CST) match or exceed research-grade actigraphy in their detection of sleep/wake states.^6,7^ Such devices can also achieve respectable sleep staging performance relative to the gold-standard, polysomnography, particularly in healthy participants without disrupted sleep.^6,7^ This augurs well for efforts to collect high-quality data about sleep patterns and interventions designed to maintain or improve them. However, there remain valid concerns about the accuracy of some CSTs, particularly when they are used in settings involving disordered or disrupted sleep that challenge accurate sleep/wake detection.^8,9^ Differing consumer, manufacturer, and professional interests and expectations would benefit from a fresh, data-informed advisory on how best to employ CST to improve sleep, health, and wellbeing.

One does not need an alarm clock with the accuracy of an atomic clock to be woken up at the right time in the morning, but such precision is necessary for GPS satellites to provide safe navigation. Similarly, it is important to recognize that the most appropriate device will differ according to its intended use and the characteristics of its user.^10^ Here, we propose three categories of uses/users requiring different levels of accuracy: (1) Those for whom accuracy is paramount, for example, clinical trials seeking to establish the efficacy of a sleep intervention; or in persons with disordered sleep patterns that may confound non-EEG based sleep detection^11^ (e.g., those with prolonged sleep latency, long periods of wake after sleep onset, or extended periods of wakefulness without movement); (2) Where moderately good accuracy is desired in persons with mostly normal sleep patterns but who may have occasional deviations (e.g., those who desire high quality sleep measurement over extended durations; large-scale, long-term population health studies on sleep patterns that involve mostly healthy sleepers; or corporate sleep health programs that monitor and reward healthy sleep patterns); (3) People or organizations who are primarily interested in tracking physical activity and are only secondarily interested in sleep, and/or those only willing to pay for basic sleep logging, akin to maintaining an automated sleep diary, will probably tolerate a lower level of accuracy in sleep assessment.

Keeping in mind these use cases, we evaluate the performance of 6 prototypical examples of commonly used sleep wearable devices^*^ (both consumer and research-grade) in healthy adults without diagnosed sleep disorders, against polysomnography measures in line with recommended guidelines.^10,12^ Devices were selected from 4 categories: (a) A research-grade dry-electrode EEG headband^†^ (∼USD 1600) that is presently available for clinical trial and academic use but is no longer a consumer product, (b) A research-grade actigraph (∼USD 500) that is primarily used to track physical activity but has also been used to measure sleep, (c) Previously evaluated high-end consumer sleep trackers (∼USD 300) whose algorithms have undergone refinement within the last 5 years and whose manufacturers have publicly documented algorithm development efforts;^13,14^ one wrist-worn and one finger-worn, and (d) Two lower-cost (< USD 60) wrist-worn wearables that to our knowledge have not undergone significant external evaluation or sleep algorithm refinement. To allow for age group comparisons to be made, participants were recruited from young (18-30y), middle-aged (31-50y) and older (51-70y) adults.

Based on the evaluation results, we make recommendations that may help researchers, clinicians, consumers, and manufacturers determine which product or development path is best for their needs.

## METHODS

### Participants and study protocol

Sixty-six adults aged 20-68 years (M(SD) = 40.2(15.7) y; 29 males; ethnically composed of Chinese: 85%, Indian: 8%, Malay: 1%, Others: 6%), consented to take part in this study. Inclusion criteria were: those who (1) habitually slept at least 5h/night (between the hours of 8 pm and 10 am), (2) had a body mass index (BMI) ≤ 35 kg/m2, (3) did not report any pre-existing sleep, neurological or psychiatric disorder, (4) did not report excessive daytime sleepiness (Epworth Sleepiness Scale scores > 10)^15^ or Berlin questionnaire scores indicating high risk of Obstructive Sleep Apnea,^16^ (5) were not on wake-promoting agents (e.g. Modafinil), stimulants (e.g. Ritalin) or sodium oxybate, (6) did not have active illness (e.g., flu), and (7) were not pregnant. Participants slept overnight in our laboratory according to their habitual bedtime and were awoken by a research assistant at their habitual wake time, if not already awake. They then completed a post-sleep questionnaire 30-60 mins after wake time to assess sleep quality and if anything disturbed their sleep during the night (Supplementary Results – Post Sleep Questionnaire). The Institutional Review Board of the National University of Singapore gave ethical approval for this work.

### Sleep measurement

#### Polysomnography (PSG)

Polysomnography (PSG) was acquired using SOMNOtouch RESP devices (SOMNOmedics GmbH, Randersacker, Germany) in a light and temperature-controlled sleep laboratory. EEG was recorded at C3 and C4 (according to the international 10–20 system), referenced to the contralateral mastoids (M2 and M1 respectively). Electro-oculography (EOG) from the right and left outer canthi, and bipolar submental electromyography (EMG1-EMG2) were also recorded. The common ground and reference electrodes were placed at Fpz and Cz, respectively. Secondary measurements also included electrocardiogram (ECG) readings and finger pulse oximetry for oxygen saturation assessment to ascertain the absence of significant sleep apnoea (Supplementary Results – Apnoea Scoring). EEG signals were sampled at a frequency of 256 Hz while impedance was kept below 10KΩ for EEG, EOG and EMG channels.

The PSG was scored from “lights off” to “lights on” times, as recorded by sleep technicians. Sleep scoring was performed using a hybrid Rechtschaffen and Kales (R&K)^17^ / American Academy of Sleep Medicine (AASM) approach.^18^ R&K stage 3/4 criteria using electrodes C3/C4 were used to define N3 sleep (following AASM guidelines). To minimize individual scorer bias, we adopted a consensus scoring approach. Three independent scoring systems were used: (1) Neurobit PSG automated scoring (Neurobit Inc., New York, USA)^19^ checked by trained lab staff, (2) Somnolyzer 24×7 automated scoring (The Siesta Group Schlafanalyse GmbH, Vienna, Austria)^20^ reviewed by experts from The Siesta Group, and (3) U-Sleep webservice automated scoring (<u>sleep.ai.ku.dk</u>) which was trained and evaluated on PSG recordings from 15,660 participants in 16 clinical studies.^21^ For each 30s epoch, a consensus score was given based on the majority score of the three scorers. If all 3 scorers differed, the scoring system of (1) was used. N1 and N2 stages were subsequently combined and labelled as ‘light sleep’ to match CST definitions, which do not distinguish between the two, while N3 was relabelled as ‘deep sleep’.

### Wearable Devices

In total, six wearable devices were evaluated in the current study with five of them worn concurrently on any one night. Two of these devices were considered research-grade wearables which also provided access to raw data: (a) Dreem 3 EEG-based headband (Beacon Biosignals, Inc.; Boston, Massachusetts, USA), and (b) Actigraph GT9X (Actigraph, Inc.; Florida, USA) accelerometer-based sensor with sampling rate set to 60Hz, while the other four were considered consumer-sleep trackers (CSTs) that utilize multi-sensor information, most commonly motion and heart rate: (c) Oura ring Gen 3, running the latest sleep staging algorithm Oura Sleep Staging Algorithm 2.0 (OSSA 2.0; Oura Health Oy, Oulu, Finland), (d) Fitbit Sense, (Fitbit, Inc., San Francisco, CA, USA), (e) Xiaomi Mi Band 7 (Xiaomi, Inc., Beijing, China), and (f) Axtro Fit3 band (Axtrosports, Inc., Singapore). All participants put on devices (a)-(d), and either the Xiaomi Mi Band 7 or the Axtro Fit3 band during the overnight session. The placement of these devices is shown in Figure 1, with the three wrist-based wearables (b, d, and e or f) placed on the non-dominant wrist in the same physical position across all participants. For brevity, we will refer to each device by its brand name (i.e., Dreem, Oura, Fitbit, Actigraph, Xiaomi, Axtro) throughout the rest of the paper.

**Figure 1.**
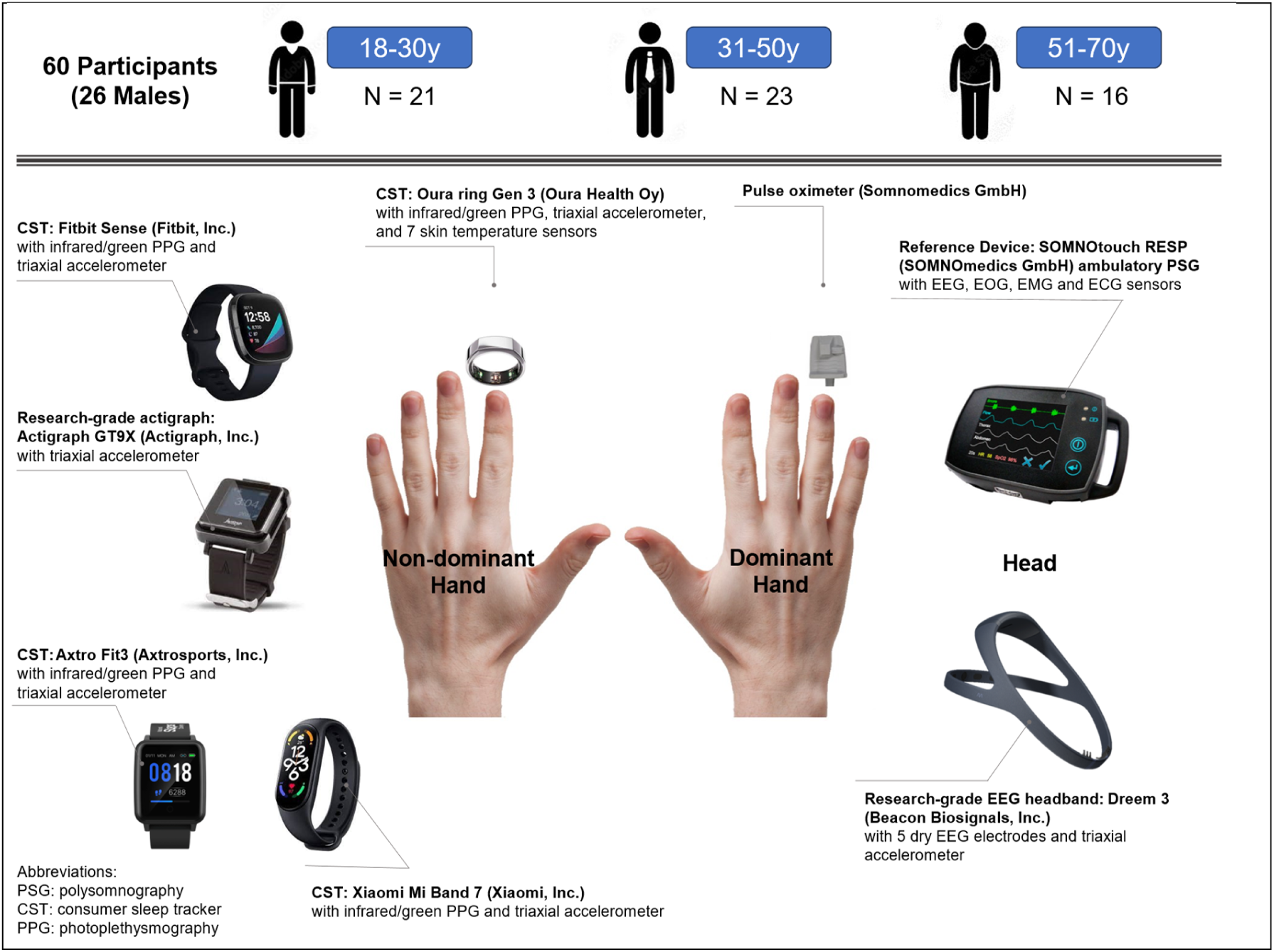
Sample demographics, device placement for a right-handed participant, devices used in the study and their sensors.

Devices were updated to the latest firmware and data were uploaded using the most recent app/software versions available, as of Mar 2023. Full details of firmware and app/software versions used are listed in Supplementary Table 1. Data from all devices except Actigraph were synced and uploaded in the morning to the respective device clouds through smartphone apps. Scored hypnograms (30s consecutive sleep staged epochs from device-determined bedtime to wake time) from each respective device vendor were then accessed through a web-based API or research portal. For Xiaomi, data were manually extracted from the smartphone app itself. Finally, for the Actigraph, triaxial accelerometer data were downloaded via a custom dock and aggregated into 1-min epochs for sleep-wake scoring using the Actilife implementation of the Cole-Kripke algorithm.^22,23^ 1-min epochs were scored from “Lights off” to “Lights on” and subsequently up-sampled to 30s epochs to match the resolution of the PSG data.

### Time synchronisation of devices

Accurate synchronization between PSG and the devices being evaluated is critical to ensure validity of the performance evaluation. At the start of each sleep session, PSG and device internal clocks were synchronized to an internet time server accessed through an internet-connected desktop computer (in the case of PSG and Actigraph) or a smartphone app (all other wearable devices).

Recording of TIB, on the Dreem and Actigraph were based on manual starts/stops (Dreem) or marked post-hoc (Actigraph) by a member of the research team. TIB was automatically detected in CSTs, based on device-specific algorithms. As such, CST hypnograms had to be adjusted to match PSG time-in-bed (TIB) based on the recorded “lights off” and “lights on” times to enable inter-device comparisons. If the TIB indicated by the device was shorter (i.e., device bedtime began after the actual lights-off time or device wake time occurred before the actual lights-on time), wake epochs were imputed to match the length of PSG TIB. Conversely, if the TIB indicated by device was longer (i.e., device bedtime began before the actual lights-off time or device wake time occurred after the actual lights-on time), device hypnograms were trimmed to match the length of PSG TIB. To determine whether this additional wake-imputation step materially affected the classification results from Oura and Fitbit, epoch-by-epoch analyses were also conducted on epochs common to each of these CSTs and PSG.

Despite clock synchronization across devices, temporal offsets between PSG and some devices were occasionally encountered with Dreem and Fitbit devices. To ensure optimal temporal alignment between PSG and the devices, and to provide the most accurate epoch-by-epoch (EBE) metrics, each device hypnogram was shifted (up to ±5 min/20 epochs) relative to the PSG.^24,25^ For each step in the shift (one epoch), correlation, accuracy, specificity, and sensitivity for sleep/wake classification were recomputed. The computation that yielded the highest value was used for subsequent analysis of that device’s data. Histograms of optimum shift values for Dreem and Fitbit are shown in Supplementary Figure 1. Note that Oura did not require any shifting (optimum shift = 0s), and that we did not perform this step for the Xiaomi or Axtro given the high classification error rate, even before the shift.

### Sleep Parameters

Sleep epochs from both PSG and wearable devices were classified into four categories: wake, light, deep, and REM sleep. Commonly reported sleep parameters, including total sleep time (TST; min of sleep between sleep onset and “lights on”), sleep onset latency (SOL; min between “lights off” to first epoch marked as sleep, regardless of sleep stage), wake after sleep onset (WASO; min awake between sleep onset and “lights on”), sleep efficiency (SE; percentage of TST while in bed from “lights off” to “lights on”) and duration spent in the different sleep stages: light sleep, deep sleep and rapid eye movement (REM) sleep, were computed. As described earlier, due to automated bed/wake time detection in the CSTs, we additionally evaluated discrepancies from these times compared to PSG marked “lights off” and “lights on” times. As Xiaomi and Axtro only begin recording at sleep onset and terminate at sleep offset (rather than bed/wake time), we compared device-recorded sleep onset/offsets in Xiaomi and Axtro to PSG-determined sleep onset and offsets.

### Missing data and partial data loss

Despite following the recommended guidelines of each device, issues with missing data and partial data loss occasionally occurred. These issues are summarized in Supplementary Table 2. Approximately one-third of participants had poor quality Dreem data, and half had either Xiaomi or Axtro data; therefore, we performed analyses in 3 subgroups to preserve power for inter-device comparisons in the largest subset of devices. The analyses were: (1) N = 60 participants with concurrent Oura, Fitbit, Actigraphy and PSG, (2) N = 40 participants with concurrent Dreem, Oura, Fitbit, Actigraphy and PSG, and (3) N = 28/20 participants with concurrent Xiaomi/Axtro, Oura, Fitbit, Actigraphy and PSG data.

## STATISTICAL ANALYSIS

### Discrepancy analyses

To visualise discrepancies between sleep measures recorded from each wearable device and PSG, Bland–Altman plots were generated using a standardized framework for performance evaluation studies.^12^ A negative bias represents underestimation by the device compared to PSG. Proportional bias, that is, how bias was affected by the magnitude of the measure, and homoscedasticity were also assessed. Bland–Altman plots demonstrating device-PSG biases for TST, SOL, WASO, SE (2-stage sleep/wake classification) and duration spent in the different sleep stages (4-stage sleep stage classification) were generated. Subgroup analyses comparing participants with high sleep efficiency (SE ≥ 85%) to those with SE of < 85% (due to either long SOL or WASO) were also performed.

To assess whether measurements from a device differed significantly from PSG, one-sample t-tests (against zero) on the device-PSG bias were conducted. In addition, to compare if biases differed across devices, separate repeated measures analysis of variance (ANOVA) for each sleep parameter with device as the within-subjects factor were also conducted. Significant interactions were followed by post-hoc paired t-tests; p-values were corrected for multiple comparisons using Bonferroni correction. We also examined whether device-PSG biases in sleep parameters differed by sex and age group. Mixed ANOVAs were employed for each sleep parameter bias as the outcome variable, with device (Dreem, Oura, Fitbit, Actigraph) as the within-subjects factor, and age group (young adults 18-30y, middle-aged 31-50y, and older adults 51-70y) or sex (male/female) as the between-subjects factor.

### Epoch by Epoch (EBE) Analysis

EBE analyses – the preferred approach to assess accuracy of binary (2-stage) and categorical (4-stage) classification^12^ were performed on 30s epochs. Sensitivity (ability of device to correctly identify ‘sleep’), specificity (ability of device to correctly identify ‘wake’), overall accuracy, and F1 score were calculated for each subject following the equations below, and then averaged across all subjects to obtain group level values.

> Sensitivity: True sleep / (False Wake + True Sleep)
>
> Specificity: True wake / (True Wake + False Sleep)
>
> Accuracy: (True Sleep + True Wake) / Total Epochs
>
> F1: True sleep / (True sleep + 0.5 x (False Sleep + False Wake))

This process was repeated for evaluating 4-stage classification performance. In addition, we also calculated Cohen’s kappa coefficient, which takes into account agreement metrics occurring by chance, and prevalence-and bias-adjusted kappa (PABAK), which adjusts for imbalances in the relative frequency of occurrence of the different sleep stage/wake epochs and bias between PSG and device metrics, using the equations below:

> Kappa = (Po-Pe)/(1-Pe)
>
> Po: Probability of agreement
>
> Pe: Probability of disagreement (chance)
>
> PABAK = (2 Pe - .05)/(1 - 0.5)=2Po-1

Kappa values are usually interpreted as follows: ≤ 0 indicates no agreement, 0.01–0.20 none to slight, 0.21–0.40 indicates fair, 0.41– 0.60 indicates moderate, 0.61–0.80 indicates substantial, and 0.81–1.00 almost perfect agreement.

In accord with the discrepancy analyses, repeated measures ANOVAs were used for EBE analyses on device-PSG agreements of accuracy, sensitivity, and specificity; followed by post-hoc paired t-tests; p-values were corrected for multiple comparisons using the Bonferroni correction.

Finally, to inspect sources of misclassification, confusion matrices were constructed. These were first generated per subject by dividing values in each cell with the corresponding marginal frequency of the reference PSG measure. Next, confusion matrices were averaged across all subjects to generate group level matrices.

Statistical analyses and data processing were performed using SPSS 27.0 (IBM Corp., Armonk, New York), MATLAB version R2017b (The Math Works, Inc., Natick, MA) and R version 4.1.1 (2021-08-10).

## RESULTS

### Performance Evaluation of High-End CST (Oura/Fitbit), Actigraphy vs. PSG (N=60)

60 participants (26 males; mean (SD) age: 38.5 (15.1) y) with acceptable concurrent Oura, Fitbit, Actigraph and PSG data contributed to the primary subgroup analysis. PSG-derived sleep measures for young (18-30y; N = 21), middle-aged (31-50y; N = 23), and older age groups (51-70y; N = 16) are presented in Table 1. While TIB and TST were similar across the groups, older adults had more WASO and less deep sleep, while young adults had longer SOL, leading to lower sleep efficiency in both groups relative to the middle-aged group.

**Table 1.** Demographic characteristics and polysomnography-determined sleep architecture of the sample. P-values <0.05 denote measures where metrics differ across age groups.

| Measure | All Subjects | Young Adults<br>18-30y<br>(N=21) | Middle-Aged<br>Adults<br>31-50y<br>(N=23) | Older Adults<br>51-70y<br>(N=16) | p-value |
| --- | --- | --- | --- | --- | --- |
| **Age** | 38.47 (15.05) | 23.19 (2.27) <sup>a,b</sup> | 37.65 (4.89) <sup>a,c</sup> | 59.69 (6.47) <sup>b,c</sup> | < .001 |
| **TIB (min)** | 433.90 (48.90) | 429.17 (57.17) | 426.28 (45.32) | 451.06 (40.02) | .12 |
| **TST (min)** | 368.88 (53.48) | 368.36 (62.81) | 374.09 (40.11) | 362.09 (59.47) | .68 |
| **WASO (min)** | 50.03 (39.32) | 37.74 (26.99) <sup>b</sup> | 43.11 (27.50) | 76.09 (54.69) <sup>a,c</sup> | < .001 |
| **SE (%)** | 85.20 (9.60) | 85.67 (8.78) <sup>b</sup> | 88.02 (6.38) <sup>c</sup> | 80.53 (12.83) <sup>b,c</sup> | .014 |
| **SOL (min)** | 14.99 (13.72) | 23.07 (18.95) <sup>a</sup> | 9.09 (4.49) <sup>a</sup> | 12.88 (9.11) | .007 |
| **Deep Sleep (min)** | 63.89 (29.85) | 83.88 (31.29) <sup>a,b</sup> | 57.83 (21.03) <sup>a</sup> | 46.38 (24.62) <sup>b</sup> | < .001 |
| **Light sleep (min)** | 227.79 (43.14) | 213.62 (46.20) | 231.48 (39.77) | 241.09 (40.79) | .16 |
| **REM Sleep (min)** | 77.20 (27.30) | 70.86 (29.41) | 84.78 (22.79) | 74.63 (29.43) | .21 |

<sup>a</sup> Young significantly different from Middle-Aged ( $p < 0.05$ ). <sup>b</sup> Young significantly different from Older Adults ( $p < 0.05$ ). <sup>c</sup> Older Adults significantly different from Middle-Aged ( $p < 0.05$ )

The following results focus on individual device-PSG agreement measures. Further details comparing performance between wearable devices are provided in Supplementary Results – Post-Hoc Between Device Comparisons.

### 2-Stage Classification Performance (Discrepancy and EBE Analyses)

Bland–Altman plots showing device-PSG biases for TST, WASO, SOL, SE and sleep-stage analyses are presented in Figures 2 and 3, respectively. Compared to PSG, none of the three devices showed significantly different TST (Table 2). Oura slightly overestimated SOL by 10.32 min, (*t =* 4.99*, p =* .002, *Cohen’s d* =.43) and underestimated WASO by 11.21 min, (*t =* 2.84*, p =* .006, *Cohen’s d* =.36). Actigraph underestimated SOL by 9.21 min, (*t =* 3.27*, p <* .001, *Cohen’s d* =.65).

**Figure 2.**
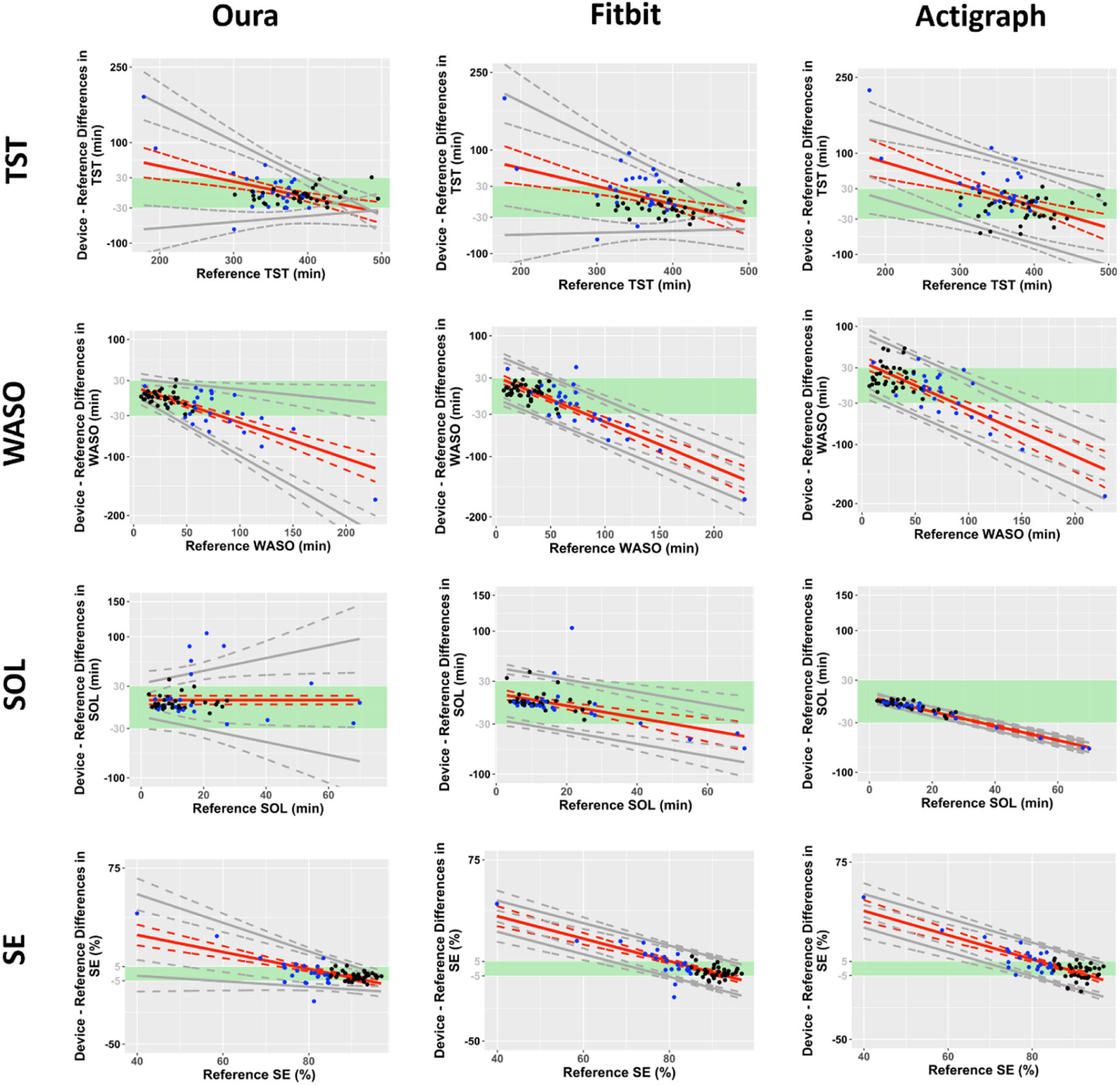
Bland–Altman plots for TST, WASO, SOL, and SE for Oura, Fitbit and Actigraph (N=60). Black dots indicate good sleepers with PSG-determined sleep efficiencies (SE) ≥ 85% while blue dots indicate poor sleepers with SE < 85%. Green boundaries indicate clinically acceptable limits of ± 30-minutes for TST, WASO and SOL or ± 5% for SE. Solid grey lines indicate 95% levels of agreement. Dotted lines indicate 95% CIs.

**Figure 3.**
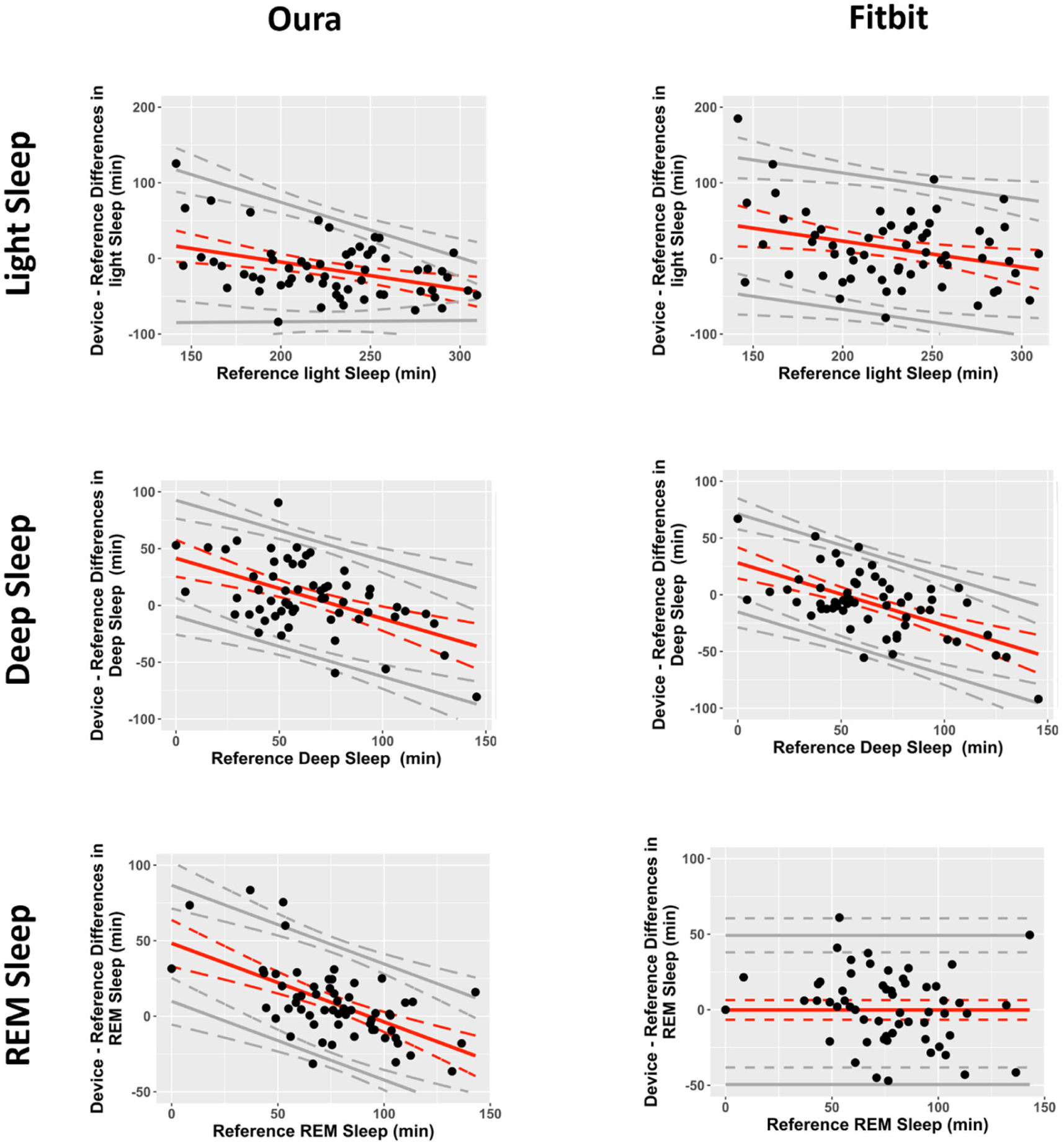
Bland–Altman plots for light sleep, deep sleep, and REM sleep for Oura and Fitbit (N=60). Solid grey lines indicate 95% levels of agreement. Dotted lines indicate 95% CIs.

**Table 2.** Discrepancy analyses comparing Oura, Fitbit and Actigraph with PSG (N=60). Values represent means and standard deviations of device-PSG biases, with positive and negative values denoting over-and underestimation compared with PSG respectively. P-values <0.05 denote measures where metrics differ across devices.

| Measure | Oura | Fitbit | Actigraph | p-value |
| --- | --- | --- | --- | --- |
| **TST (min)** | .89 (34.56)<sup>b</sup> | 5.97 (40.66) | 8.99 (44.11)<sup>b</sup> | .042 |
| **SE (%)** | .16 (8.23)<sup>b</sup> | 1.16 (9.24) | 1.96 (10.18)<sup>b</sup> | .061 |
| **SOL (min)** | 10.32 (24.03)<sup>** a,b</sup> | -.43 (21.91)<sup>a,c</sup> | -9.22 (14.31)<sup>*** b,c</sup> | <.001 |
| **WASO (min)** | -11.21 (30.56)<sup>** a,b</sup> | -5.54 (34.68)<sup>a,c</sup> | .23 (40.37)<sup>b,c</sup> | <.001 |
| **Light (min)** | -14.74 (38.41)<sup>** a</sup> | 13.34 (48.31)<sup>* a</sup> | N.A. | <.001 |
| **Deep (min)** | 7.55 (30.50)<sup>a</sup> | -7.25 (27.54)<sup>* a</sup> | N.A. | <.001 |
| **REM (min)** | 8.08 (24.21)<sup>* a</sup> | -.13 (25.19)<sup>a</sup> | N.A. | .008 |
| **Bedtime (min)** | 14.28 (25.85)<sup>*** a</sup> | 3.17 (25.89)<sup>* a</sup> | N.A. | .002 |
| **Wake time (min)** | 2.65 (5.98)<sup>***</sup> | 1.06 (11.09) | N.A. | .331 |

Asterisks indicate whether bias is significantly different from zero (one-sample t-test with Bonferroni correction for multiple comparisons): \*p < 0.05; \*\*p < 0.01; \*\*\*p < 0.001
Letters indicate significant differences in bias between devices: <sup>a</sup> Oura significantly different from Fitbit (p < 0.05). <sup>b</sup> Oura significantly different from Actigraph (p < 0.05). <sup>c</sup> Fitbit significantly different from Actigraph (p < 0.05).

In contrast to the non-significant biases in TST, standard deviations were wide (> 34 min) and limits of agreement were proportionally larger for poor sleepers with low SE (< 85%, i.e. those with longer SOL and/or WASO). When considering only good sleepers with PSG-determined sleep efficiencies ≥ 85%, the majority of the datapoints for TST, SOL and WASO were within clinically acceptable limits of ± 30 mins (blue points within green shaded areas in Figure 2 and blue points in Supplementary Figure 2) for Oura (> 97.14% of points), Fitbit (> 80% of points) and Actigraph (> 82.85% of points). The same was observed for SE bias with a majority of datapoints within clinically acceptable limits of ± 5%; Oura (> 82.86% of points), Fitbit (> 77.14% of points) and Actigraph (> 57.14% of points). Examples of well aligned hypnograms from good sleepers are shown in Supplementary Figure 8 (e.g., participant ID: 3,4,17), with one exemplar showcased in Figure 4a.

**Figure 4.**
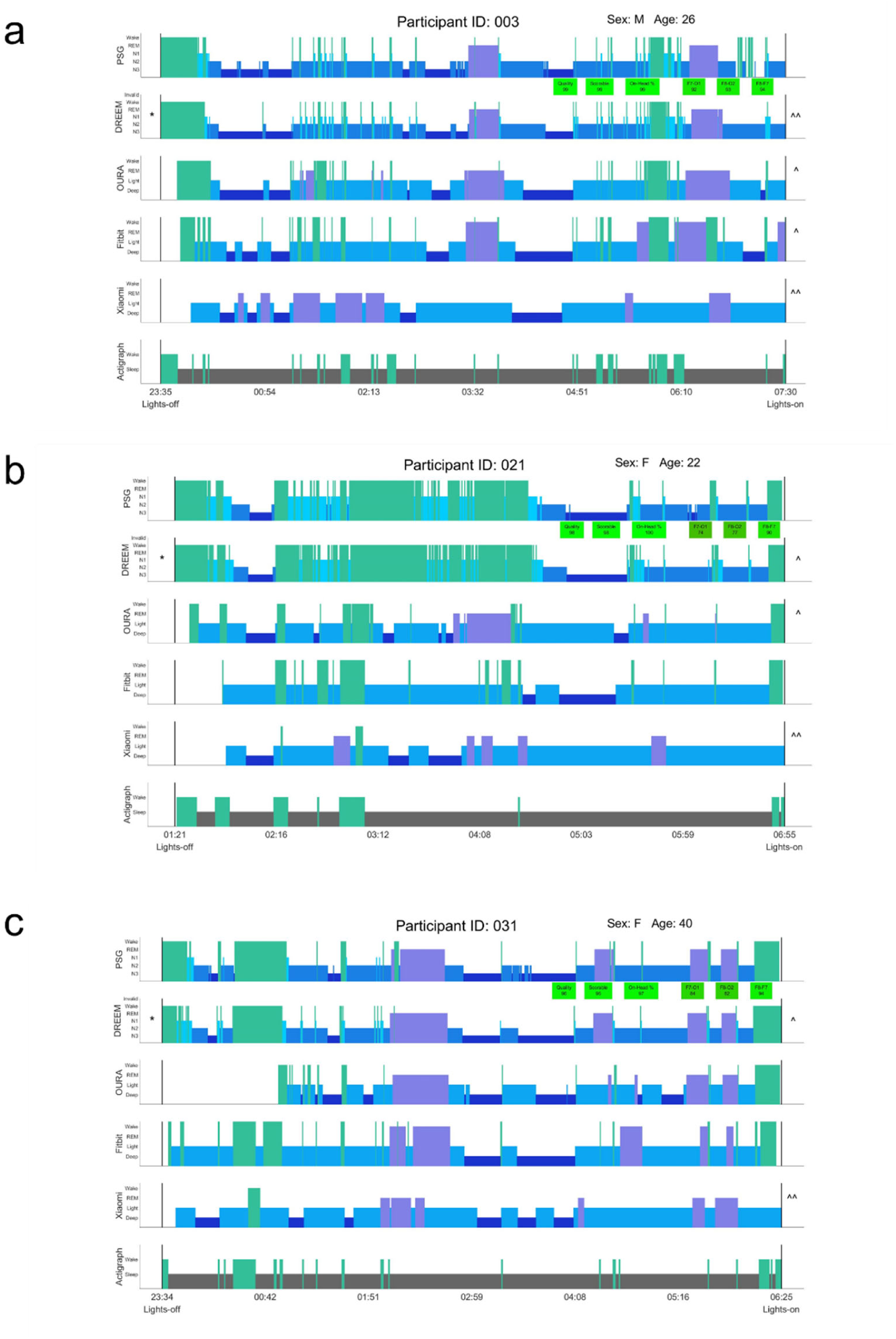
Examples of well and poorly aligned wearable hypnograms compared to PSG. **(a)** Well-aligned hypnogram from a good sleeper with SE ≥ 85%. Apart from Xiaomi, sleep-wake and sleep-stage epochs largely aligned with PSG. **(b)** Participant with a long mid-sleep WASO period but who appeared still while awake based on the absence of motion in Actigraph. Only the Dreem headband which has EEG sensors accurately detected this prolonged wake episode. **(c)** Delayed bedtime detection in Oura, resulting in overestimation of SOL due to wake imputation from “lights off”, and underestimation of WASO as the earlier PSG-defined WASO epochs would be classified as SOL rather than WASO.

With Fitbit and Actigraph, poor sleepers also had significantly higher device-PSG discrepancies for TST, WASO, and SE compared with good sleepers (*ts ≥* 3.34, *ps* < .002, *Cohen’s ds* ≥ .98), while for Oura this difference only reached statistical significance for WASO, (*t =* 3.27*, p =* .003, *Cohen’s d* =.99); with comparable performance for TST and SE (*ts <* 1.72, *ps ≥* .097).

For EBE analyses, although overall accuracies were between 87-91%, devices were better at detecting sleep (sensitivity values: 93-95%) than wake (specificity values: 56-73%; Table 3). This was evident when inspecting the hypnograms of participants who were lying awake with little movement in the middle of the sleep period - the CSTs and motion-based Actigraph severely underestimated wake. Examples of such cases are shown in Supplementary Figure 8 (e.g., participant ID: 21, 65, 67), with one exemplar showcased in Figure 4b.

**Table 3.**
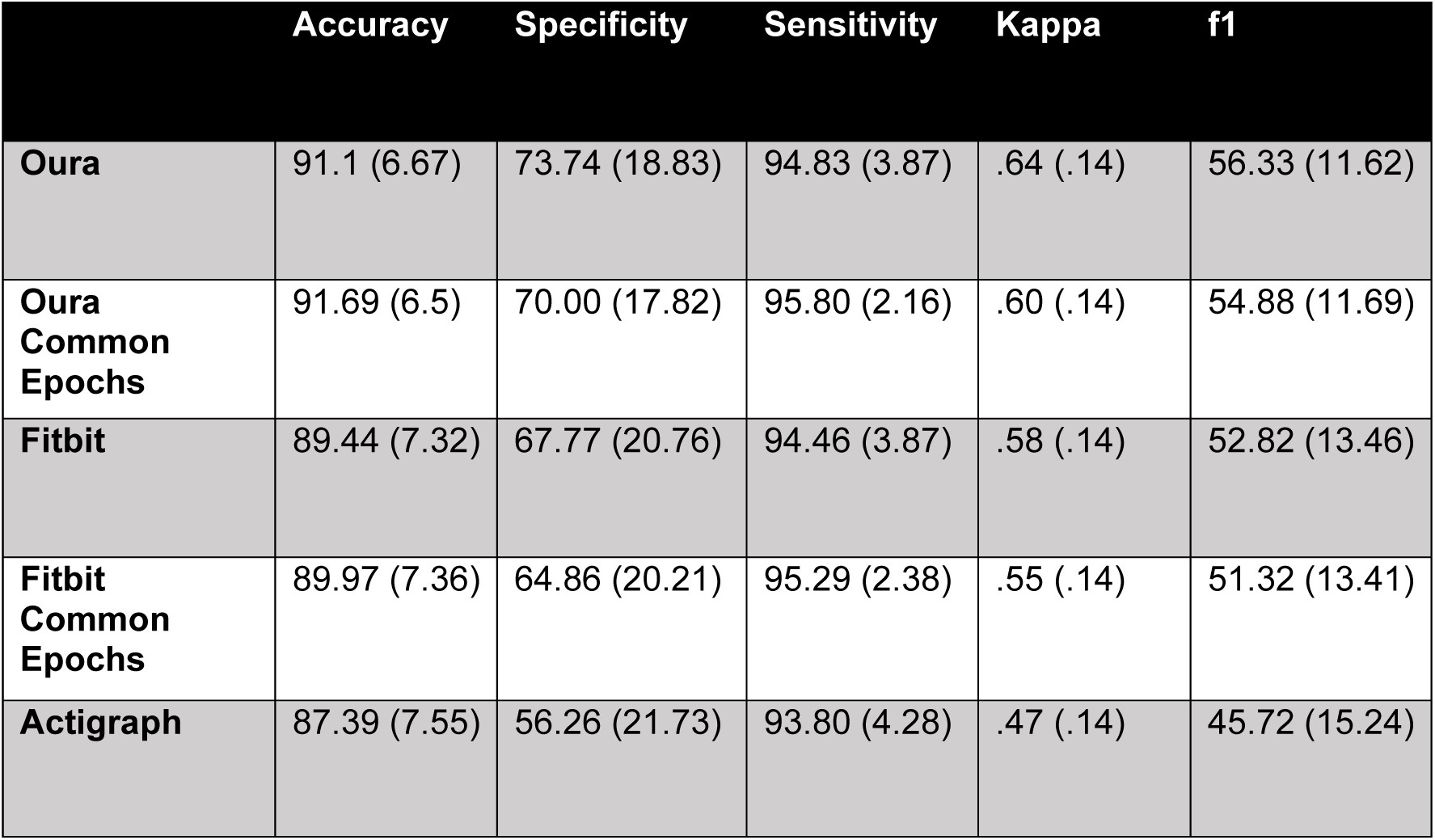
Epoch-by-epoch (EBE) analyses comparing 2-stage sleep/wake agreement between Oura, Fitbit and Actigraph with PSG (N=60). Common-epoch analyses refer to analyses constrained to epochs that were present in both CST and PSG (i.e., before wake imputation in CST devices).

|  | Accuracy | Specificity | Sensitivity | Kappa | f1 |
| --- | --- | --- | --- | --- | --- |
| **Oura** | 91.1 (6.67) | 73.74 (18.83) | 94.83 (3.87) | .64 (.14) | 56.33 (11.62) |
| **Oura**<br>**Common**<br>**Epochs** | 91.69 (6.5) | 70.00 (17.82) | 95.80 (2.16) | .60 (.14) | 54.88 (11.69) |
| **Fitbit** | 89.44 (7.32) | 67.77 (20.76) | 94.46 (3.87) | .58 (.14) | 52.82 (13.46) |
| **Fitbit**<br>**Common**<br>**Epochs** | 89.97 (7.36) | 64.86 (20.21) | 95.29 (2.38) | .55 (.14) | 51.32 (13.41) |
| **Actigraph** | 87.39 (7.55) | 56.26 (21.73) | 93.80 (4.28) | .47 (.14) | 45.72 (15.24) |

Of the non-EEG based wearables, Oura showed significantly better 2-stage classification performance with accuracy of 91.1%, kappa of .64, and PABAK of .82 compared to Fitbit with 89.44% accuracy, kappa of .58, and PABAK of .79, (ts > 5.68, ps < .001) and Actigraph with 87.39% accuracy, kappa of .47, and PABAK of .75, (ts > 6.90, ps < .001).

### 4-Stage Classification Performance (Discrepancy and EBE Analyses)

Light sleep was underestimated by Oura by 14.74 min and overestimated by Fitbit by 13.34 min (Figure 3 and Table 2). Conversely, deep sleep was overestimated by Oura by 7.55 min (not significant), while Fitbit underestimated it by 7.25 min. For REM sleep, only Oura was significantly different to PSG, where it was underestimated by 8.08 min.

For EBE analyses, confusion matrices show that REM sleep classification was the best with Oura (82% correctly classified) while the other stages were slightly less accurate (74-76% correctly classified). For Fitbit, light sleep was the most accurately classified stage (77%) while deep sleep classification was the least accurate at only 57%, which was misclassified as light sleep 41% of the time (Supplementary Figure 3). Oura had a 4-stage kappa range of .55-.70 compared with Fitbit with .45-.60 kappa values.

### Automated Bedtime and Wake Time Detection on CSTs

As bed and wake times were automatically determined by the CSTs, we compared their deviations from “lights off” and “lights on” times marked in the PSG (Table 2).

Oura appeared to have a more conservative estimation of bedtime with significantly delayed bedtime detection compared with Fitbit by 11.1 min, (*t =* 3.22*, p* = .002, *Cohen’s d* =.42, Table 2 and Supplementary Figure 4), requiring a longer consolidated immobile period to initiate detection of a sleep period. This could have affected the discrepancy and EBE analyses independently of sleep/wake classification performance, resulting in overestimation of SOL due to wake imputation from “lights off”, and conversely, underestimation of WASO if there were occurrences of PSG-defined WASO before device-determined bedtimes. Examples of such cases are shown in Supplementary Figure 8 (e.g., participant ID: 11, 16, 31, 40), with one exemplar showcased in Figure 4c. In addition, 51/60 and 54/60 of points lay within ± 30 mins of “lights off” for Oura and Fitbit respectively (Supplementary Figure 4).

Wake time biases were smaller (1-3 min on average) with no significant difference between Oura and Fitbit (*t =* .98*, p* = .33). 59/60 and 58/60 of points were within ± 30 mins of “lights on” for Oura and Fitbit respectively.

### Performance Evaluation of Dreem compared with High-End CSTs (Oura / Fitbit), Actigraphy and PSG (N=40)

The second subgroup analysis comprised 40 participants (22 males; mean (SD) age: 38.03 y (14.74)) who had usable Dreem, Oura, Fitbit, Actigraph and PSG data. Sample sizes for the young, middle-aged and older age groups were N = 13, 19 and 8 respectively.

### 2-Stage Classification Performance (Discrepancy and EBE Analyses)

Dreem showed, numerically, the lowest TST, SOL, WASO and SE discrepancies with PSG compared to Oura, Fitbit, and Actigraph, along with smaller standard deviations (∼12-15 min) and narrower limits of agreement (Table 4 and Figure 5). However, repeated measures ANOVA on sleep measurement biases only showed a significant main effect of device for SOL, where Dreem significantly outperformed Oura and Actigraph (SOL: *F* = 9.34, *p <* .001, ηp^2^ = .19). Even on nights with highly fragmented sleep, Dreem was able to identify wake periods with high accuracy even when the participant appeared to be lying awake with little movement (Figure 4b).

**Figure 5.**
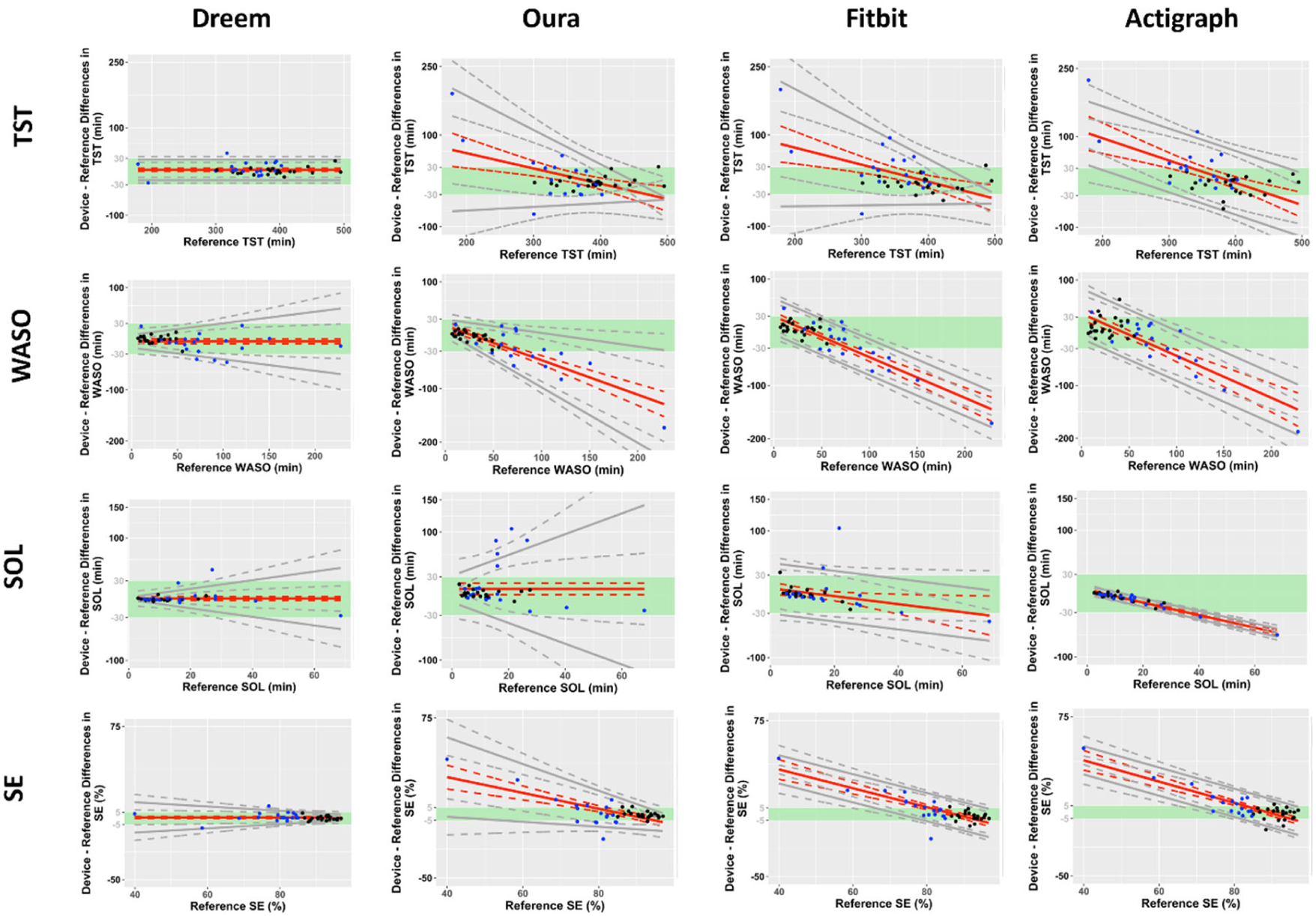
Bland–Altman plots for TST, WASO, SOL, and SE for Dreem, Oura, Fitbit and Actigraph (N=40). Black dots indicate good sleepers with PSG-determined sleep efficiencies (SE) ≥ 85% while blue dots indicate poor sleepers with SE < 85%. Green boundaries indicate clinically acceptable limits of ± 30-minute bias duration for TST, WASO and SOL or ± 5% for SE. Solid grey lines indicate 95% levels of agreement. Dotted lines indicate 95% CIs.

**Table 4.** Discrepancy analyses comparing Dreem, Oura, Fitbit and Actigraph with PSG (N=40). Values represent means and standard deviations of device-PSG biases, with positive and negative values denoting over-and underestimation compared with PSG respectively. P-values <0.05 denote measures where metrics differ across devices based on repeated measures ANOVAs.

| Measure | Dreem | Oura | Fitbit | Actigraph | p-value |
| --- | --- | --- | --- | --- | --- |
| **TST (min)** | 3.85 (12.01) * | 5.08 (40.31) | 10.16 (44.31) | 14.51 (47.18) # | .21 |
| **SE (%)** | .77 (2.82) | 1.17 (9.62) | 2.18 (10.12) | 3.41 (10.85) # | .20 |
| **SOL (min)** | 1.03 (10.04) | 11.10 (28.12) * | .51 (21.77) | -8.24 (12.44) *** | <.001 |
| **WASO (min)** | -4.88 (15.16) * | -16.18 (34.33) ** | -10.68 (38.75) | -6.28 (43.64) | .11 |
| **Light (min)** | -19.80 (21.70) *** | -15.68 (40.72) * | 16.48 (50.10) * |  | <.001 |
| **Deep (min)** | 16.16 (17.79) *** | 10.65 (25.09) * | -4.29 (25.24) |  | <.001 |
| **REM (min)** | 7.49 (15.05) ** | 10.10 (25.41) * | -2.03 (24.38) |  | .013 |

Asterisks indicate whether bias is significantly different from zero (one-sample t-test with Bonferroni correction for multiple comparisons).
Multiple comparison corrected p-values: \*p < 0.05; \*\*p < 0.01; \*\*\*p < 0.001; # p < .06

For EBE analyses, Dreem also outperformed the other wearables, with overall accuracy of 95.02%, sensitivity of 97.34%, specificity of 78.21%, kappa of .76, and PABAK of .9 (Table 5).

**Table 5.**
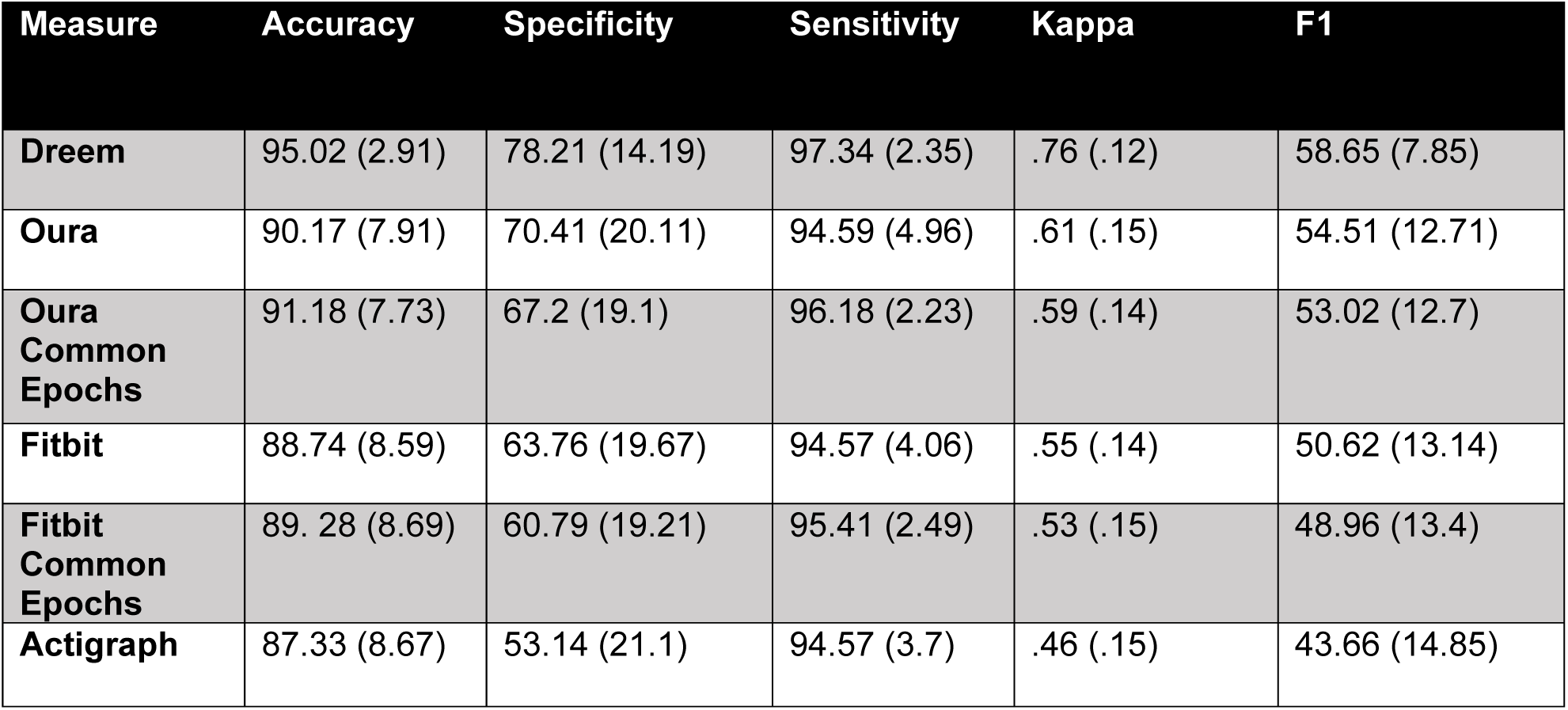
Epoch-by-epoch (EBE) analyses comparing 2-stage sleep/wake agreement between Dreem, Oura, Fitbit and Actigraph with PSG (N=40). Common-epoch analyses refer to analyses constrained to epochs that were present in both CST and PSG (i.e., before wake imputation in CST devices).

### 4-Stage Classification Performance (Discrepancy and EBE Analyses)

Dreem significantly underestimated light sleep by 19.8 min and overestimated deep and REM sleep by 16.16 min and 7.49 min respectively. However, limits of agreement were much smaller with Dreem than with any CST (Table 4 and Figure 6).

**Figure 6.**
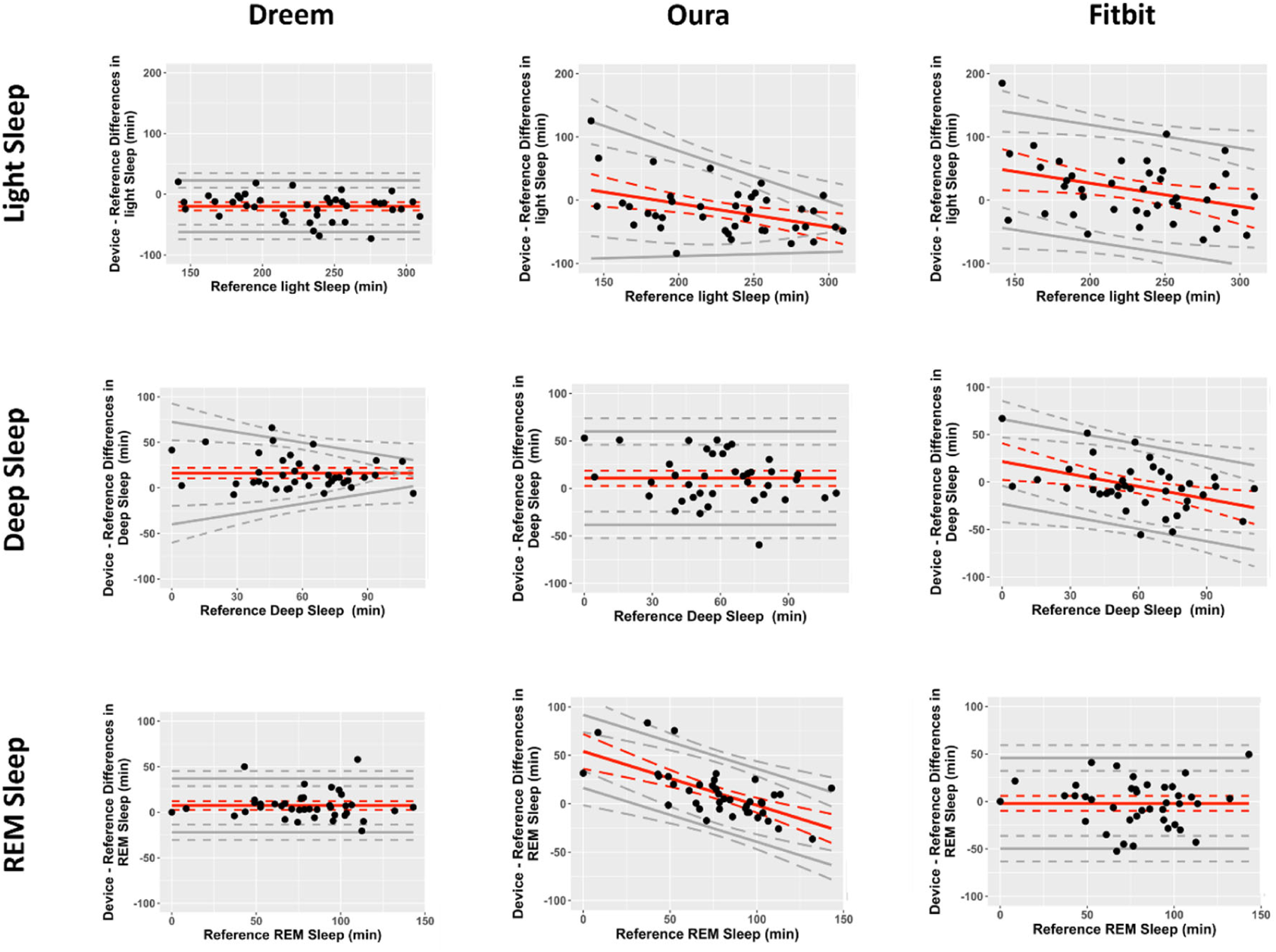
Bland–Altman plots for light sleep, deep sleep, and REM sleep for Dreem, Oura, and Fitbit (N=40). Solid grey lines indicate 95% levels of agreement. Dotted lines indicate 95% CIs.

For EBE analyses, Dreem again led 4-stage classification performance with 84%, 94% and 93% classification accuracy for light, deep and REM sleep, respectively (Supplementary Figure 5). Even brief stage transitions were accurately detected leading to superior performance of Dreem compared with the non-EEG based CSTs (examples of such cases are shown in Supplementary Figure 8; e.g., participant ID: 5 and 8). Overall, only Dreem and Oura achieved kappa values indicating substantial agreement with PSG (kappas ≥ .60).

### Age and Sex Influences on Sleep Tracking Accuracy (N=40)

#### 2-Stage Classification Performance

Repeated measures ANOVA on device-PSG biases showed a significant age group by device interaction for TST (*F* = 4.48, p < .001, ηp^2^ = .20), SE (*F* = 3.78, *p* = .002, ηp^2^ = .17), and WASO (*F* = 5.54, *p* < .001, ηp^2^ = .23). Biases for TST, SE and WASO were largest in the oldest age group (51-70y; *F* > 6.56, *p* < .004, ηp^2^ = .26); discrepancies across age groups were smallest with Dreem (Figure 7a). Of the non-EEG based devices, discrepancies were more homogenous with Oura than with Fitbit and Actigraph, specifically for TST and SE biases in the older adults (*ts* > 3.55, *p*s < .009). Similar trends were observed in the primary subgroup analyses with N = 60 participants (Supplementary Figure 6).

**Figure 7.**
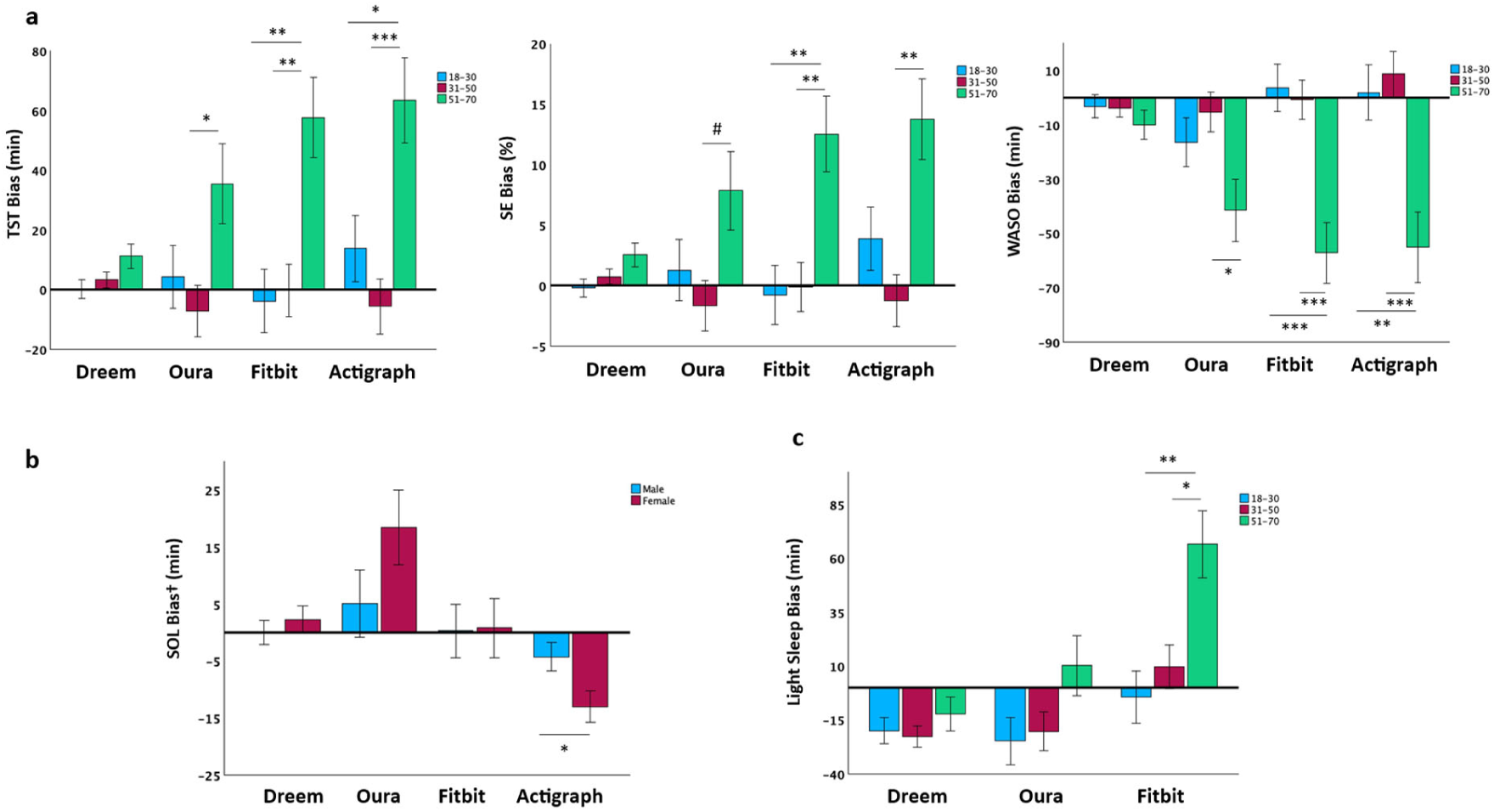
Age and sex influences on sleep tracking accuracy. 2-stage classification: **(a)** Significant device by age group interactions were observed for TST, SE and WASO bias, whereby Dreem outperformed other devices across age groups, followed by Oura, particularly in older adults. **(b)** For sex by device bias interactions, only SOL bias was significant, whereby Actigraph tended to underestimate SOL more in females vs. males. **4-stage classification: (c)** A significant device by age group interaction was present only for light sleep bias; where Fitbit tended to overestimate this more in the older compared to the younger age group. Multiple comparison corrected p-values: *p < 0.05; **p < 0.01; ***p < 0.001; # p = .055. ^†^: SOL calculated from PSG “lights off” to detected sleep onset. Blue, red, and green bars refer to young (18-30y), middle-aged (31-50y), and older age groups (51-70y), respectively.

For sex by device bias interactions, only SOL bias was significant, where Actigraph tended to underestimate SOL more in females vs. males (*F* = 4.48, *p* < .001, ηp^2^ = .196, (Figure 7b).

#### 4-Stage Classification Performance

A significant device by age group interaction was present only for light sleep bias, (*F* = 3.31, *p* = .015, ηp^2^ = .15) which Fitbit overestimated more in the older compared to the younger age group (Figure 7c).

### Performance Evaluation of Low-Cost Consumer-Based Devices (Xiaomi / Axtro) compared with High-End CSTs (Oura / Fitbit), Actigraphy and PSG

The third subgroup analysis was performed with N=28 (Xiaomi) and N=20 (Axtro) participants respectively, after removal of unusable records.

Both low-cost consumer-based devices significantly underperformed Oura, Fitbit, and Actigraph for multiple sleep measurements across both 2-stage and 4-stage classification metrics. Device-PSG biases were significantly higher for TST, WASO, SE, as well as light and REM sleep for Xiaomi (*ts* > 3.03, *p*s < .005, *Cohen’s d* > .57) and significantly higher for WASO, SOL, as well as light, deep, and REM sleep for Axtro, (ts > 2.52, ps < .019, *Cohen’s d* > .51, compared to the other CST devices and Actigraph (Supplementary Table 3,4 and Supplementary Figure 7a).

EBE analyses of these low-cost devices showed similarly poor performance. While 2-stage classification accuracy for sleep detection (sensitivity) was high (94-95%), these devices were very poor at identifying wake (specificity, 33%), resulting in overall low kappa scores (< .31). 4-stage classification metrics were also poor, with kappa scores < .33 (Supplementary Figure 7b).

Comparisons between device-recorded sleep onset/offsets to PSG-determined sleep onset and offsets however showed that Xiaomi provided acceptable estimates, with 25/28 (89%) of points lying within ± 30 mins of sleep onset, and 27/28 (96%) of points within ± 30 mins of sleep offset. For Axtro, although the average mean discrepancy appeared to be just a few mins, only 12/20 (60%) of points lay within ± 30 mins of sleep onset, and 16/20 (80%) of points within ± 30 mins of sleep offset.

## DISCUSSION

Performance evaluation of 6 wearable sleep trackers across 4 representative device categories supports a stratified approach to selecting a device that integrates technological, research and clinical considerations. We first discuss our findings in relation to 2-stage sleep/wake classification as there is a wealth of epidemiologic and actigraphy data relating sleep duration and sleep disruption (WASO) to indicators of health, wellbeing, and mortality.^27–29^ Our evaluations show that device performance was significantly affected by sleep efficiency; this was particularly notable for lower quality devices that have poor wake specificity. Therefore, it is useful to consider good and bad sleepers separately when evaluating performance. We then discuss choices for demanding situations involving users who do not have normative nocturnal sleep patterns and users who desire accurate 4-category sleep staging.

### CST and 2-Stage Sleep Tracking

The largest group of CST users are mainly healthy working age adults between 37-55 years of age^30^ who are mid to higher SES individuals keen on maintaining or improving sleep health.^31^ On nights with good sleep (SE ≥ 85%), a majority of the datapoints for TST, SOL and WASO were within clinically acceptable limits of ± 30 mins, particularly for the Oura ring, with >97% of points meeting this arbitrary threshold generally accepted by clinicians.^32–35^ This result is important for proper longitudinal assessment of sleep variability, its effects on outcomes of interest, for evaluating the effectiveness of an intervention, or fluctuations in the severity of disordered sleep.

The superior results of the higher quality CSTs relative to the research-grade actigraph^36^ attests to the value of adding heart rate (HR) variability detection to actigraphy in CST^13,14,37^ together with better training data and improved algorithms. A review found that later Fitbit models incorporating HR sensing/sleep-staging surpassed earlier accelerometery-only devices (no HR models: sensitivity: 87-99%, specificity: 10-52%, HR models: sensitivity: 95-96%, specificity: 58-69%),^38^ and also research-grade actigraphy (GT3X: sensitivity: 90-95%, specificity: 35-46%),^36,39^ particularly for specificity measures. Importantly, merely additional sensors alone does not assure improvement in measurement precision. This is illustrated by the poor sleep-wake classification performance of the low-cost devices (Xiaomi and Axtro) that incorporate PPG HR detection but are without documented efforts to improve sleep assessment and/or proper quality evaluations.

Consumers should critically appraise claims of ‘high accuracy’ during sleep-wake agreement testing conducted by comparing epoch-by-epoch 2-stage classification with PSG. As sleep efficiency is over 80-85% in most healthy people, even an inaccurate wearable can achieve ‘accuracy’ and ‘sensitivity’ for sleep detection of ≥ 90% simply by assigning ‘sleep’ as the default stage. In contrast, ‘specificity’ for identifying wake is the more discriminating metric. It ranged from 33% for the low-cost CSTs to 53% for Actigraph, 62% for Fitbit, 70% for Oura and 78% for Dreem.

Specificity is important for identifying fragmented sleep, and high values are difficult to achieve when periods of WASO are short and frequent, as in those suffering from sleep disorders.^40^ The low-cost wearables we assessed failed to adequately identify wake periods even in relatively healthy older adults. Our results are similar to a recent Xiaomi (Mi Band 5) evaluation study, where specificity was 38% and kappa was .27 in participants without sleep disorders.^41^ Such devices would be inadequate for population health studies or clinical applications unless all that is required is to identify when an individual went to sleep and woke up.^42^ For this purpose, the Xiaomi device may be deemed fit-for-use as it showed that the majority, 25/28 observations were within ± 30 mins of PSG determined sleep onset, while 27/28 points were within ± 30 mins of sleep offset.

The kappa score, which reflects overall agreement between a device and PSG, is another useful measure of wearable performance as it accounts for agreement metrics occurring by chance. The rank order of kappa scores of the devices we tested followed that of the specificity results. Only the Dreem headband and Oura ring achieved kappa values indicating substantial agreement (≥ .60), while kappa scores for Xiaomi and Axtro were poor (< .31).

By analyzing the hypnograms of 60 participants and viewing concurrent data on 5 devices at a time, we observed that short bouts of wake or sleep tended to be ‘smoothed’ or ignored to different extents by each wearable. This likely contributed to the heterogeneity of specificity and kappa results. For example, when a person does not transition directly into consolidated sleep but fluctuates between wakefulness and light sleep over the course of minutes, bedtime detection may be delayed until sleep is more consolidated. This affects SOL and TST measures non-uniformly across different devices. During lab testing, and with Dreem/Actigraph, bedtime (lights out) and wake time (lights on) timings are user-triggered/marked by researchers. However with most CSTs, bedtime and wake time detections are automated such that sleep staging only commences when consolidated sleep is deemed to have started and stops when significant and/or prolonged activity is detected.^24^ Oura tended to report later sleep onset^‡^, after sleep was more clearly consolidated compared to Fitbit, and it also terminated sleep early if there was a long WASO period (e.g., 30 min), while Fitbit would tend to concatenate such separate bouts of sleep and classify them as a single main sleep period.

The quality of sleep assessment showed an interaction between age and device. As expected from age-related changes in sleep, the biases for TST and WASO were largest in older participants (51-70y) who tended to have poorer sleep. Unsurprisingly, the EEG-based Dreem headband had the lowest discrepancy for these measures. Of the non-EEG devices, discrepancies were also more homogenous with Oura than with Fitbit and Actigraph. This could reflect a greater sample size and age diversity in the training data used to develop Oura’s latest sleep staging algorithm OSSA 2.0, which was trained on 326 adolescents andadults across 7 independent datasets and 757 nights of polysomnography recording^§^. This was larger than the training set for an earlier algorithm we evaluated in 2022^43^, and allowed for feature normalization that adjusts for inter-individual differences in physiology (e.g., nocturnal HR that tends to be higher in older than younger adults).^14^ By way of comparison, the Actigraph’s Cole-Kripke algorithm was refined using data from only 32 men and 9 women, 23 of whom had sleep or psychiatric disorders.^44^

Consistent with a very large-scale survey of sleep patterns in the US, UK, and the Netherlands^45^, our study found edge cases in which young adults had low sleep efficiency (and duration) in conjunction with prolonged sleep latency, whereas in the edge cases involving older adults, the issue was with sleep continuity and/or earlier awakenings. When sleep disturbances were artificially induced in a laboratory, they also affected sleep tracking performance, prolonging SOL and lowering SE.^6^ Overall, the finding that sleep assessment is less accurate and more variable in older persons (Figure 7) for non-EEG based systems, even at the 2-stage sleep/wake classification is contributed to by inherent issues with specificity (wake detection) described earlier. It remains to be seen if market growth among older users can drive development of methods to transcend the challenges posed in accurately detecting motionless wakefulness in older adults.

## 4-Stage Sleep Classification Performance

Although consumers, clinicians and some researchers pay close attention to 4-stage (wake, light sleep, deep sleep, and REM) classification performance, these results should only be considered after assessment of ‘traditional’ actigraphy measures because they depend on accurate 2-stage, sleep/wake classification. Unsurprisingly, the EEG-based Dreem headband led 4-stage performance with 85%, 94% and 93% classification accuracy for light, deep and REM sleep respectively. Dreem would be the first choice for studying people with disordered sleep, for applications such as highly sensitive clinical trials or the evaluation of sleep interventions with projected small effect sizes, as its high wake detection specificity circumvents inaccuracies when assessing participants with low SE. However, the headband was not tolerated by at least 25% of participants in our study (38% of records had to be excluded for poor quality recordings or when the device was off-head for more than 10% of the time), must be manually activated to start a recording and has a more complex data uploading procedure compared to the non-EEG based CSTs evaluated here. Another caveat with Dreem is that for unknown reasons, the temporal alignment of Dreem-staged epochs could be offset by as much as 5 min. To obtain the best EBE statistics in comparison with PSG, each Dreem recording had to be temporally shifted. Dreem also tended to slightly underestimate wake, often misclassifying it as N1/N2.^46^

Specificity values for specific sleep stages were overall higher than sensitivity values (except for light sleep on Fitbit), particularly for deep sleep. This is because deep sleep epochs are the fewest among the different sleep stages, and individual misclassifications carry more weight for sensitivity than specificity measures.^47^ Of the higher quality CSTs, REM sleep classification was the best with Oura (82% correctly classified) while other stages were only slightly less accurate (74-75% correctly classified). For Fitbit, light sleep was the most accurately classified stage (77%) while deep sleep classification only averaged 57%, mirroring previous findings in healthy adults (light: 76-81%, deep: 49-53%, REM: 69-74%).^6,48^ The lower cost CSTs did not have acceptable 4-stage classification performance (kappa < .33) and, therefore, should not be used for any use-cases requiring 4-stage classification.

### Recommendations for Different Use Cases

Our evaluations only involved off-the shelf measurements from tested devices. This is the most likely scenario for wearable users or sleep scientists operating outside of major research groups who have privileged access to raw sensor data, the means to collect and to process these signals, machine learning expertise and/or the resources to evaluate optimized methodology against PSG. Without modifications, the EEG-based Dreem headband (or its equivalent) is the best system when sleep measurement accuracy is paramount, but price, user comfort and convenience may be significant concerns.

For the second category encompassing most researchers and critical consumers in whom sleep is mostly normal and who desire high quality longitudinal sleep measurement, a well-validated tracker such as the Oura ring or Fitbit (or their equivalents) is the best device of choice. In addition to sleep and activity measurement, these devices also come with built-in trend tracking features that provide weekly/monthly summaries as well as sleep hygiene tips and even digital sleep coaching. Oura additionally allows for firmware locking and blocking of feedback from users, features important for longitudinal research/observational studies. While the use of a research-grade actigraph like the GT9X provides acceptable sleep tracking as an adjunct to physical activity monitoring in research settings, the incremental value of having raw accelerometry data without HR sensors may be overestimated. Further, as recently demonstrated,^23^ the equivalence of sleep measurements from research-grade actigraphy cannot be assumed.

Finally, for the third group of users who may only require a lower-cost tracker to help log sleep periods over the long-term and not require accurate 2-stage or 4-stage classification, the Xiaomi or its equivalent could be a reasonable cost-effective alternative. Such devices could serve as reminders to workers who have irregular sleep to improve habits and can be deployed on a scale not possible with more expensive devices. Over time, as sleep measurement methodology, societal valuation of sleep and pricing models evolve, the quality of these devices is likely to improve.

### Strengths and Limitations

We tested 5 devices concurrently to provide direct comparisons across key wearable device categories in three equally sized groups of participants of different ages to ensure that older participants with typically lower sleep efficiency were included in the assessment.

Inter-rater reliability of PSG metrics typically only averages 80-82%^49^ and is lower when assessing people with sleep disorders.^50,51^ This is important to remember when making comparisons between PSG and non-EEG based wearables as the reference instrument is in fact, imperfect as well.^52^ To increase the likelihood that the PSG reference was as reliable as possible, we used consensus-based scoring to reduce the likelihood of scorer bias affecting the overall results, combining both ML-based and trusted human reader approaches.

While the primary aim of the present work was to validate 2-stage and 4-stage classification performance across commonly used sleep trackers, limitations of automatic bedtime/sleep period detection constrain performance of algorithms in consumer devices (Oura, Fitbit, Xiaomi, Axtro), relative to Dreem and Actigraph, where sleep periods are manually started or marked in the recordings. For example, in one participant, sleep was so fragmented that no sleep periods were detected, and as such there was no data for the sleep algorithm to stage. This automated sleep period detection could also lead to much shorter SOL/WASO durations displayed on the smartphone app than those computed from “lights off” markers as done in the present work, particularly if sleep periods are only initiated once consolidated sleep is detected. However, when considering only classification of epochs within the detected sleep period (without imputed wake intervals), specificity and sensitivity measures of Oura and Fitbit were still better than Actigraph. Some consumer devices allow the user to manually start and stop sleep periods, or to edit them later, but care should also be taken to ensure that these bed/wake time markers are interpreted consistently across users (e.g., ‘time I got into bed’ vs. ‘time I intended to sleep’.)

A lab-based protocol, while well-controlled may not reflect sleep behaviors in the real world^6^ ; for example, engagement with pre-and post-sleep activities (e.g. texting/reading in bed) is common with pervasive use of smartphones and tablet devices.^53^ As such, future work will need to evaluate the performance of these devices in settings that reflect real-world sleep behavior as well as collect contextual information that would enable meaningful interpretation of the data output.^54^

## CONCLUSION

In sum, we provided a thorough evaluation of 4 categories of wearable sleep technology in a good number of participants across different ages, using a comprehensive array of test measures and have made pragmatic suggestions for device class purchases based on different user requirements to balance currently deliverable performance with cost considerations.

## Data Availability

Summarized data underlying the results presented in the study are available upon reasonable request.

## ACKNOWLEDGEMENTS

We are grateful to Yahsmit Lepcha, Liang Tian and Nicha Turton for their assistance in data collection. This work was supported by funds from Oura Health Oy, the Yong Loo Lin School of Medicine, Lee Foundation, and the National Medical Research Council Singapore (STaR May2019-001) awarded to Dr Michael W.L. Chee.

## AUTHOR CONTRIBUTIONS

J.O., A.W. and M.C. conceptualized and designed the study. K.W. and N.C. coordinated the recruitment of participants and data collection. K.W., S.G. and H.G. performed the data analysis, and contributed to the first draft of the paper. J.O., A.W. and M.C provided critical input for analysis and reviewed the paper critically. J.O., H.G. and M.C wrote the final version of the paper. All authors have approved the final submitted version of the paper.

## Footnotes

* We use the term ‘wearable devices’ to collectively refer to both research and consumer-grade trackers throughout this manuscript. However, when describing performance of individual categories of devices, we refer to these as either research-grade (Dreem, Actigraph) or consumer-grade (CST) trackers (Oura, Fitbit, Xiaomi, Axtro).

† As of late August 2023, Dreem was acquired by Beacon Biosignals and it was announced that it will switch to a subscription model with differential pricing for clinical trial and academic partners.

‡ The positive bias in the Bland-Altman plots for SOL is often related to an artefact of how SOL is defined in lab-based studies. Delayed detection of sleep onset results in a longer interval between lights off and detected sleep. In fact, device measured SOL (as reported on the smartphone app) is typically shorter relative to what is determined by PSG.

§ Numbers derived from direct communication with Oura.

