## Supplementary Files for "A Data Driven Approach for Choosing a Wearable Sleep Tracker"

**SUPPLEMENTARY RESULTS**

**Post-Sleep Questionnaire**

A post-sleep questionnaire was completed by participants on a smartphone within 1h after "lights on". This questionnaire comprised 11-items that probed subjective sleep quality, mood and sleepiness on a Likert scale and 2 open-ended questions describing the difficulties they faced (if any) trying to fall asleep or if anything made them uncomfortable during the night. Mean (SD) of ratings by participants included in the final analyses (N=60) are shown in the table below:

| Question | Mean (SD) |
|---|---|
| How would you rate the quality of your sleep? (1 – good, 5 – poor) | 2.98 (0.95) |
| Compared to usual, how was the quality of your sleep? (1 – much better, 5 – much worse) | 3.53 (0.87) |
| How rested or refreshed did you feel when you woke up for the day? (1 – very well rested, 5 – not rested at all) | 3.00 (0.92) |
| Compared to usual, how rested or refreshed do you feel? (1 – much better, 5 – much worse) | 3.30 (0.79) |
| Compared to usual, how is your mood today? (1 – much better, 5 – much worse) | 2.93 (0.41) |
| How tired are you? (1 – not at all, 4 – extremely) | 1.88 (0.69) |
| How sleepy are you? (1 – not at all, 4 – extremely) | 1.82 (0.68) |
| How alert are you? (1 – not at all, 4 – extremely) | 2.75 (0.65) |
| How does this compare to a usual night of sleep at home? (1 – much better, 5 – much worse) | 3.43 (0.81) |
| How long did take for you to fall asleep (in minutes)? | 38.33 (42.41) |
| Compared to usual, was this… (1 – shorter, 2 – about the same, 3 – longer) | 2.58 (0.62) |
| Did you have difficulty falling asleep last night? (1 – yes, 2 – no) | 1.37 (0.49) |

Open-ended responses (N=60) were manually categorised as: 1) environmental disturbances, 2) general equipment disturbances, and 3) Dreem-specific disturbances. Twenty-one (35%) participants indicated trouble sleeping due to environmental issues (e.g., too quiet, temperature too low, bed), 24 (40%) indicated disturbances due to equipment in general, while 15 (25%) participants specifically indicated that the Dreem band interfered with their sleep.

**Apnoea Scoring**

AHI scoring was performed with the DOMINO software packaged with the SOMNOmedics PSG system. Oxygen desaturation criterion was set to lower than 3% below baseline lasting for at least 8s. For the final sample analysed (N=60), mean (SD) of the AHI scores was 6.32 (6.03), in the mild range.

**Post-hoc Between-Device Comparisons**

**Performance Evaluation of Mid-Range CST (Oura / Fitbit), Actigraphy vs. PSG (N=60)**

- **2-Stage Classification Performance (Discrepancy and EBE Analyses)**

Repeated measures ANOVA on observed device-PSG biases for TST, WASO, and SOL showed a significant main effect of the device on all three metrics (TST: F = 3.25, p = .042, $\eta p^2 = .05$; WASO: F = 7.61, p < .001, $\eta p^2 = .11$; SOL: F = 20.42, p < .001, $\eta p^2 = .25$ Table 2, Figure 2). Post-hoc paired t-tests demonstrated that Oura had significantly less TST overestimation compared with Actigraph by an average of 8.1 min (t = 2.32, p =.023, Cohen's d = .30); a similar trend was observed with Fitbit where there was 5.08 min less TST overestimation but this was not statistically significant, (t = 1.66, p = .103).

For WASO, Actigraph significantly outperformed both Oura and Fitbit with an average of 11.44 min (t = 3.52, p < .001, Cohen's d =.45), and 5.78 min (t = 2.09, p = .041, Cohen's d = .27) less underestimation, respectively. Fitbit significantly performed better than Oura with 5.67 min less underestimation of WASO (t = 2.06, p = .044, Cohen's d = .26). For SOL bias we observed significantly better performance of Fitbit, with an average of 10.74 min less overestimation (t = 3.45, p < .001, Cohen's d = .45), and 8.62 min less underestimation (t = 3.76, p < .001, Cohen's d = .49) compared with Oura and Actigraph, respectively. Oura and Actigraph performed significantly differently in the opposite direction, (t = 5.43, p < .001, Cohen's d =.70); while Oura significantly overestimated SOL by 10.32 (24.03) min, Actigraph significantly underestimated it by 9.22 (14.31) min, Table 2, Figure 2.

For EBE analyses, repeated measures ANOVA on accuracy, sensitivity and specificity measures indicated a main effect of device (accuracy: F = 49.09, p < .001, $\eta$p2 = .45; sensitivity: F = 7.69, p < .001, $\eta p^2 = .12$; specificity: F = 18.93, p < .001, $\eta p^2 = .24$, Table 3). Post hoc paired t-tests showed significantly higher accuracy, sensitivity, as well as specificity values of Oura compared to Fitbit and Actigraph (ts ≥ 1.99, ps < 0.05, Cohen's ds ≥ .26). Similarly, Fitbit outperformed Actigraph across all EBE metrics (ts ≥ 2.21, Ps < 0.031, Cohen's ds ≥ .28). When analyses were constrained to epochs common to each of these devices and PSG, the CST classification performance was still significantly better than the Actigraph, (ts ≥ 1.99, ps < 0.05, Cohen's ds ≥ .26).

- **4-Stage Classification Performance (Discrepancy and EBE Analyses)**

Oura and Fitbit significantly differed to each other in opposite directions for both light (t = 5.91, p < .001, Cohen's d =.76) and deep sleep biases (t = 4.76, p < .001, Cohen's d =.62). For REM sleep, Fitbit outperformed Oura with significantly less overestimation by 8.2 min (t = 2.74, p = .008, Cohen's d =.35).

For EBE analyses, repeated measures ANOVA on accuracy, sensitivity and specificity measures indicated a main effect of device, (accuracy: F = 32.31, p < .001, $\eta$p2 = .35; sensitivity: F = 3.99, p = .05, $\eta p^2 = .06$; specificity: F = 9.87, p = .003, $\eta p^2 = .14$); indicating better performance of Oura compared with other devices.

**Performance Evaluation of Dreem compared with Mid-Range CST (Oura / Fitbit), Actigraphy and PSG (N=40)**

- **2-Stage Classification Performance (Discrepancy and EBE Analyses)**

Repeated measures ANOVA on observed device-PSG biases showed a significant main effect of device only for SOL, whereby Dreem significantly outperformed Oura and Actigraph (SOL: F = 9.34, p < .001, $\eta p^2 = .19$, Table 4, Figure 5).

For EBE analyses, repeated measures ANOVA on device-PSG agreements of accuracy, sensitivity and specificity indicated a significant main effect of device on all three metrics considered (accuracy: F = 28.24, p < .001, $\eta p^2 = .42$; sensitivity: F = 9.67, p < .001, $\eta$p2 = .19; specificity: F = 21.35, p < .001, $\eta p^2 = .35$, Table 5). Post hoc paired t-tests showed significantly higher accuracy, sensitivity, as well as specificity values of Dreem compared to other devices (ts ≥ 2.16, ps < 0.037, Cohen's ds ≥ .34). For the other devices we observed similar trends to the N=60 sample analyses with Oura significantly preforming better than Actigraph across agreement measures (ts ≥ 2.85, ps < 0.007, Cohen's ds ≥ .24), and outperforming Fitbit in accuracy and specificity (ts ≥ 2.88, ps < 0.028, Cohen's ds ≥ .36). Likewise, Fitbit outperformed Actigraph in accuracy and specificity (ts ≥ 3.17, ps = 0.003, Cohen's ds ≥ .50).

- **4-Stage Classification Performance (Discrepancy and EBE Analyses)**

Repeated measures ANOVA on observed device-PSG biases for sleep staging measurements showed a significant main effect of the device for light sleep, deep sleep and REM sleep bias (light sleep: F = 18.26, p < .001, $\eta p^2$ = .32; deep sleep: F = 12.53, p < .001, $\eta$p2 = .24; REM sleep: F = 4.58, p = .013, $\eta$p2 = .11; Table 4, Figure 6). Compared with Fitbit, Dreem and Oura performed significantly differently and in opposite directions for both light and deep sleep estimation. While light sleep was significantly underestimated by Dreem and Oura, it was significantly overestimated by Fitbit (t > 4.85, p < .001, Cohen's d > .77). Conversely, Dreem and Oura significantly overestimated deep sleep, while Fitbit underestimated it, (t > 3.66, p < .001, Cohen's d > .58). However, Dreem did not show proportional bias, unlike Oura and Fitbit as non-EEG devices that tended to give readings showing regression to the population mean in the form of negative proportional bias. For REM sleep bias Fitbit outperformed both Dreem and Oura with significantly less overestimation, (t > 2.38, p < .022, Cohen's d > .38).

For EBE analyses, repeated measures ANOVA on accuracy, sensitivity and specificity measures indicated a main effect of device, (accuracy: F = 18.86, p < .001, $\eta$p2 = .33; sensitivity: F = 7.12, p = .001, $\eta p^2$ = .15; specificity: F = 14.46, p < .001, $\eta p^2$ = .27); indicating better 4-stage classification performance of Dreem compared with other devices, followed by Oura.

### SUPPLEMENTARY TABLES

**Supplementary Table 1** Firmware and app/software versions used in the study.

| Wearable Device | Firmware Versions | App/Software Versions |
|---|---|---|
| Dreem 3 | 5.7.15 | 1.12.10 |
| Oura Ring Gen3 | 2.8.60-2.8.61 | Oura 4.9.3-4.10.3 |
| Fitbit Sense | 44.128.6.17 | 3.82.fitbit-mobile-38251031-533196116 |
| Actigraph GT9X | 1.7.2 | Actilife 6.13.4 |
| Xiaomi Mi Band 7 | 2.0.0.2 | Zepp Life 6.7.1 |
| Axtro Fit3 | 1.0.3 | hiSG+ 1.8.4 |

**Supplementary Table 2** Reasons for excluded records.

a) Excluded records due to missing data.

| Wearable Device | Reason | Number of Records |
|---|---|---|
| Dreem 3 | - | - |
| Oura Ring Gen3 | Highly fragmented sleep throughout the night, resulting in no sleep period initiated. | 1 |
| Fitbit Sense | Device malfunction. | 1 |
| Actigraph GT9X | Technician error. | 1 |
| Xiaomi Mi Band 7 | - | - |
| Axtro Fit3 | Delayed data sync. | 3 |

b) Excluded records due to partial/poor quality data.

| Wearable Device | Reason | Number of Records |
|---|---|---|
| Dreem 3 | Dreem quality metric < 70 and off-head metric > 10%. | 23 |
| Oura Ring Gen3 | Early termination of sleep period due to long WASO. | 1 |
| Fitbit Sense | Split sleep periods due to long WASO, periods with <3h did not contain sleep stages*.<br><br>No sleep stages recorded* despite single sleep period detected, due to <3h limitation (1) or loose band (1). | 2<br><br>2 |
| Actigraph GT9X | | |
| Xiaomi Mi Band 7 | | |
| Axtro Fit3 | Split sleep periods due to long WASO, periods with <3h did not contain sleep stages. | 8 |

* Only Fitbit ‘Classic’ stages (Wake, Restless and Sleep) were recorded.

**Supplementary Table 3** Discrepancy analyses comparing Xiaomi, Oura, Fitbit, and Actigraph with PSG (N=28).

| Measure | Oura | Fitbit | Xiaomi | Actigraph |
|---|---|---|---|---|
| **TST (min)** | -1.23 (28.61) | 3.34 (31.46) | 24.92 (31.07)*** | 12.45 (33.73)* |
| **SE (%)** | -0.29 (7.67) | 0.62 (7.57) | 5.75 (7.50)*** | 2.96 (8.32) |
| **SOL (min)** | 11.38 (25.87)* | -2.79 (29.12) | -4.27 (18.42) | -13.96 (18.11)*** |
| **WASO (min)** | -10.14 (22.59)* | -.55 (26.05) | -20.66 (27.50)*** | 1.52 (31.14) |
| **Light (min)** | -12.66 (34.92) | 9.80 (51.45) | 43.27 (50.02)*** | |
| **Deep (min)** | 1.38 (27.56) | -11.91 (29.59)* | 2.98 (33.28) | |
| **REM (min)** | 10.05 (20.47)* | 5.45 (23.56)* | -21.32 (32.87)** | |

Significant biases using one-sample t-test against zero.
Multiple comparison corrected p-values: *$p < 0.05$; **$p < 0.01$; ***$p < 0.001$

**Supplementary Table 4** Discrepancy analyses comparing Axtrofit, Oura, Fitbit, and Actigraph with PSG (N=20).

| Measure | Oura | Fitbit | Axtrofit | Actigraph |
|---|---|---|---|---|
| **TST (min)** | 7.31 (43.44) | 14.46 (50.51) | 14.02 (81.03) | 9.96 (59.24) |
| **SE (%)** | 1.57 (9.63) | 2.97 (11.11) | 2.26 (19.17) | 1.86 (13.22) |
| **SOL (min)** | 8.04 (25.08) | -1.90 (10.36) | 38.52 (56.82) ** | -5.50 (8.48)** |
| **WASO (min)** | -15.35 (40.66) | -12.56 (44.00) | -52.54 (50.74) *** | -4.46 (53.86) |
| **Light (min)** | -16.02 (43.61) | 20.29 (54.92) | -21.69 (49.94) * | |
| **Deep (min)** | 17.12 (35.01) * | 1.31 (28.38) | 80.23 (39.49) *** | |
| **REM (min)** | 6.21 (22.94) | -7.15 (25.36) | -44.52 (24.27) *** | |

Significant biases using one-sample t-test against zero.
Multiple comparison corrected p-values: *$p < 0.05$; **$p < 0.01$; ***$p < 0.001$

### SUPPLEMENTARY FIGURES

**Supplementary Figure 1.** Histograms of optimum hypnogram shift values within a ±5 min (300 seconds) window for the Dreem and Fitbit, based on maximum sleep-wake classification accuracy compared to gold-standard PSG.

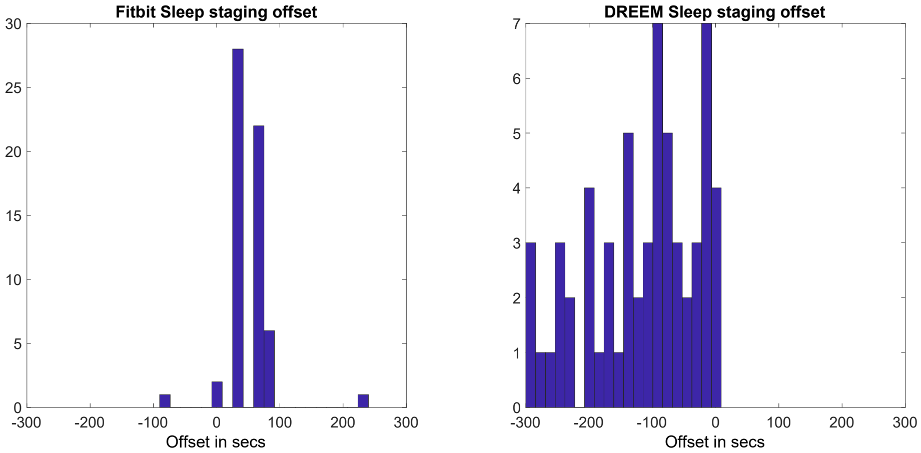


**Supplementary Figure 2.** Bland–Altman plots for TST, WASO, and SOL for Oura and Fitbit (FB) categorised by sleep efficiencies (SE ≥ 85% depicted by black dots vs SE < 85% depicted by blue dots).

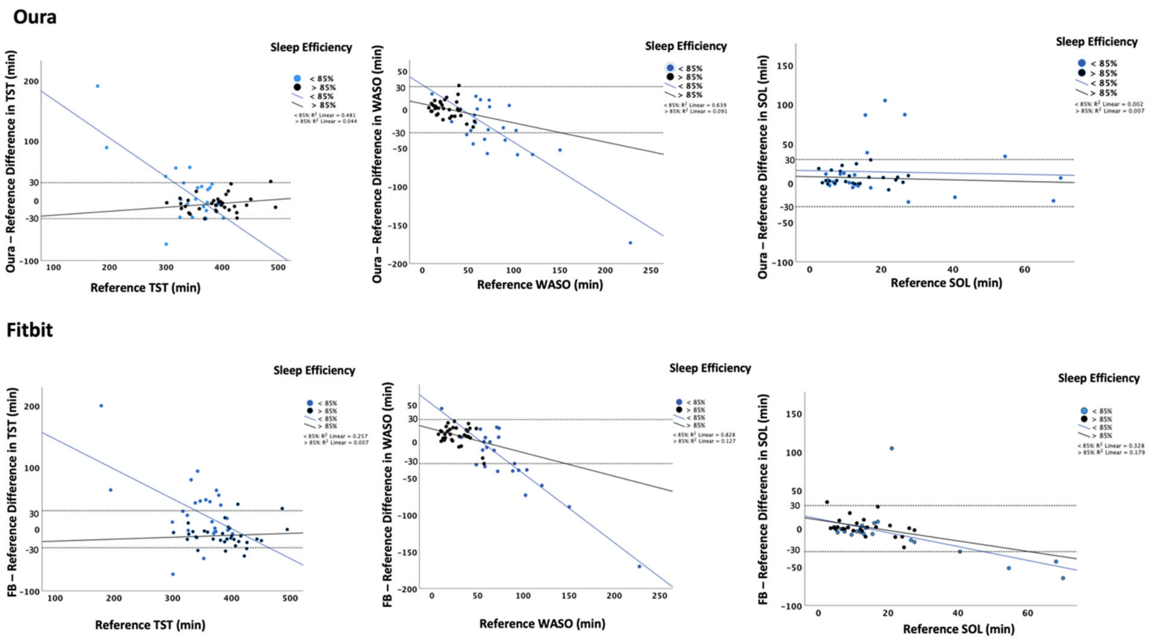

**Supplementary Figure 3.** Confusion matrices for 4-stage classification accuracy for Oura and Fitbit (N=60).

**Oura**

| Reference | Device wake | Device Light | Device Deep | Device REM |
|---|---|---|---|---|
| **Wake** | 0.74 (0.19) | 0.18 (0.14) | 0.02 (0.03) | 0.07 (0.08) |
| **Light** | 0.08 (0.06) | 0.76 (0.08) | 0.1 (0.08) | 0.07 (0.05) |
| **Deep** | 0.01 (0.03) | 0.26 (0.23) | 0.74 (0.23) | 0 (0) |
| **REM** | 0.02 (0.03) | 0.15 (0.12) | 0.01 (0.02) | 0.82 (0.14) |

**Fitbit**

| Reference | Device wake | Device Light | Device Deep | Device REM |
|---|---|---|---|---|
| **Wake** | 0.68 (0.21) | 0.25 (0.18) | 0.01 (0.02) | 0.06 (0.07) |
| **Light** | 0.08 (0.06) | 0.77 (0.10) | 0.07 (0.06) | 0.08 (0.06) |
| **Deep** | 0.01 (0.01) | 0.41 (0.26) | 0.57 (0.27) | 0.01 (0.03) |
| **REM** | 0.04 (0.06) | 0.27 (0.26) | 0.02 (0.04) | 0.68 (0.28) |

**Supplementary Figure 4.** Bland Altman plots for bed/wake time detection (Oura/Fitbit).

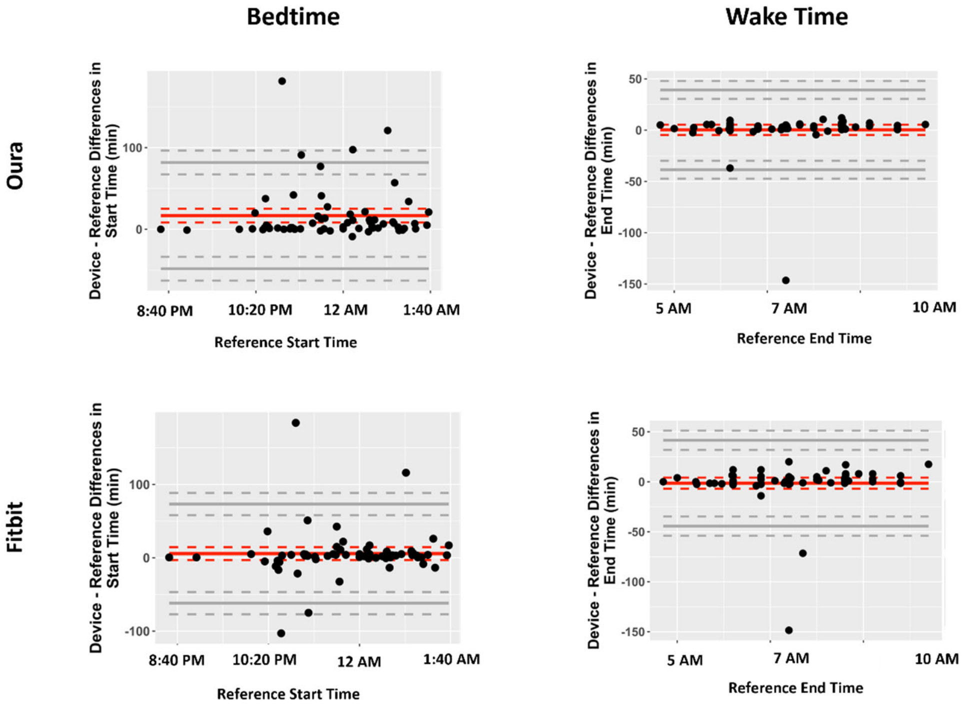

**Supplementary Figure 5.** Confusion matrices for 4-stage classification accuracy for Dreem, Oura and Fitbit (N=40).

**Dreem**

| Reference | Device wake | Device Light | Device Deep | Device REM |
|---|---|---|---|---|
| **Wake** | 0.78 (0.14) | 0.16 (0.11) | 0.01 (0.02) | 0.05 (0.07) |
| **Light** | 0.03 (0.03) | 0.84 (0.07) | 0.08 (0.07) | 0.04 (0.04) |
| **Deep** | 0 (0.01) | 0.06 (0.07) | 0.94 (0.07) | 0 (0) |
| **REM** | 0.02 (0.03) | 0.05 (0.05) | 0 (0) | 0.93 (0.06) |

**Oura**

| Reference | Device wake | Device Light | Device Deep | Device REM |
|---|---|---|---|---|
| **Wake** | 0.7 (0.2) | 0.19 (0.15) | 0.02 (0.04) | 0.08 (0.1) |
| **Light** | 0.07 (0.07) | 0.75 (0.08) | 0.1 (0.07) | 0.07 (0.06) |
| **Deep** | 0.01 (0.03) | 0.25 (0.25) | 0.74 (0.25) | 0 (0) |
| **REM** | 0.02 (0.03) | 0.15 (0.12) | 0 (0.01) | 0.82 (0.14) |

**Fitbit**

| Reference | Device wake | Device Light | Device Deep | Device REM |
|---|---|---|---|---|
| **Wake** | 0.61 (0.19) | 0.3 (0.17) | 0.01 (0.02) | 0.08 (0.08) |
| **Light** | 0.06 (0.03) | 0.79 (0.10) | 0.08 (0.06) | 0.07 (0.06) |
| **Deep** | 0.01 (0.01) | 0.43 (0.29) | 0.55 (0.29) | 0.01 (0.03) |
| **REM** | 0.04 (0.07) | 0.30 (0.28) | 0.02 (0.05) | 0.64 (0.31) |

**Supplementary Figure 6.** Device-PSG discrepancies by (a) age and (b) sex (N=60). Only significant device by age and device by sex interactions are shown. Multiple comparison corrected p-values: *p < 0.05; **p < 0.01; # p = .059

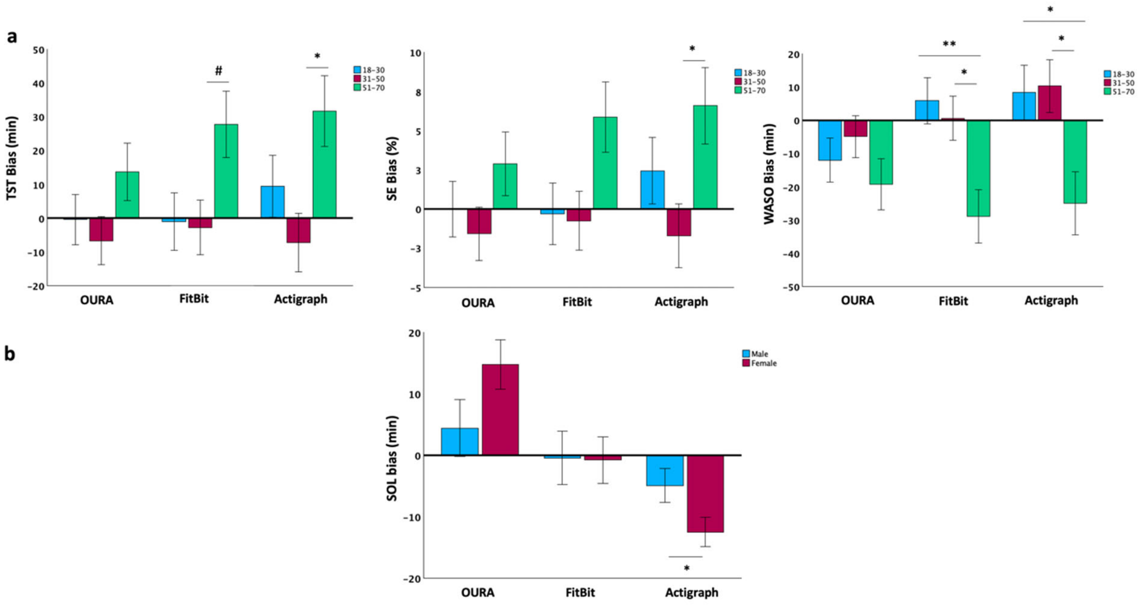

**Supplementary Figure 7.** (a) Bland-Altman plots for Xiaomi/Axtro. (b) Confusion matrices for 4-stage classification accuracy for Xiaomi/Axtro

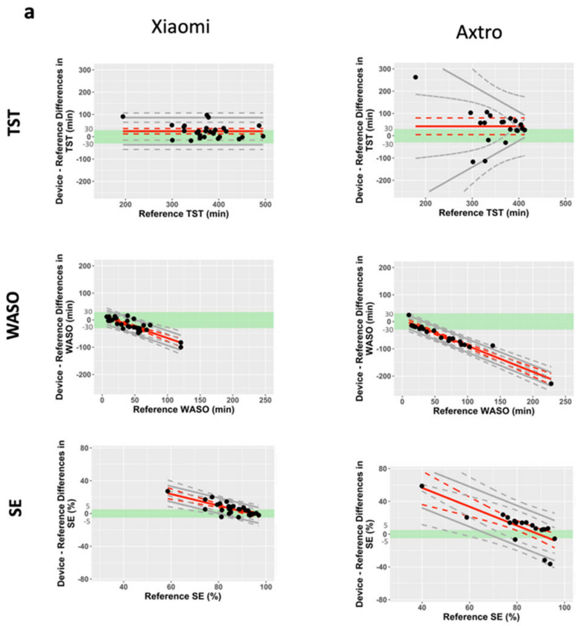


**b**

Xiaomi

| reference | device_wake | device_light | device_deep | device_REM |
|---|---|---|---|---|
| wake | 0.33 (0.2) | 0.53 (0.19) | 0.07 (0.1) | 0.07 (0.08) |
| light | 0.06 (0.06) | 0.67 (0.12) | 0.15 (0.07) | 0.11 (0.06) |
| deep | 0 (0.02) | 0.44 (0.17) | 0.46 (0.18) | 0.1 (0.11) |
| REM | 0.04 (0.06) | 0.69 (0.18) | 0.05 (0.07) | 0.22 (0.16) |

Axtro

| reference | device_wake | device_light | device_deep | device_REM |
|---|---|---|---|---|
| wake | 0.33 (0.28) | 0.43 (0.20) | 0.13 (0.12) | 0.10 (0.09) |
| light | 0.06 (0.11) | 0.53 (0.13) | 0.33 (0.12) | 0.09 (0.04) |
| deep | 0.06 (0.21) | 0.32 (0.20) | 0.59 (0.23) | 0.03 (0.6) |
| REM | 0.02 (0.05) | 0.50 (0.22) | 0.40 (0.23) | 0.08 (0.07) |

**Supplementary Figure 8.** Stacked hypnograms from 5 concurrent devices (PSG, Dreem, Oura, Fitbit, Xiaomi/Axtro and Actigraph) across 66 participants, from “lights off” to “lights on". Participant IDs, age and sex are also shown at the top of each plot.

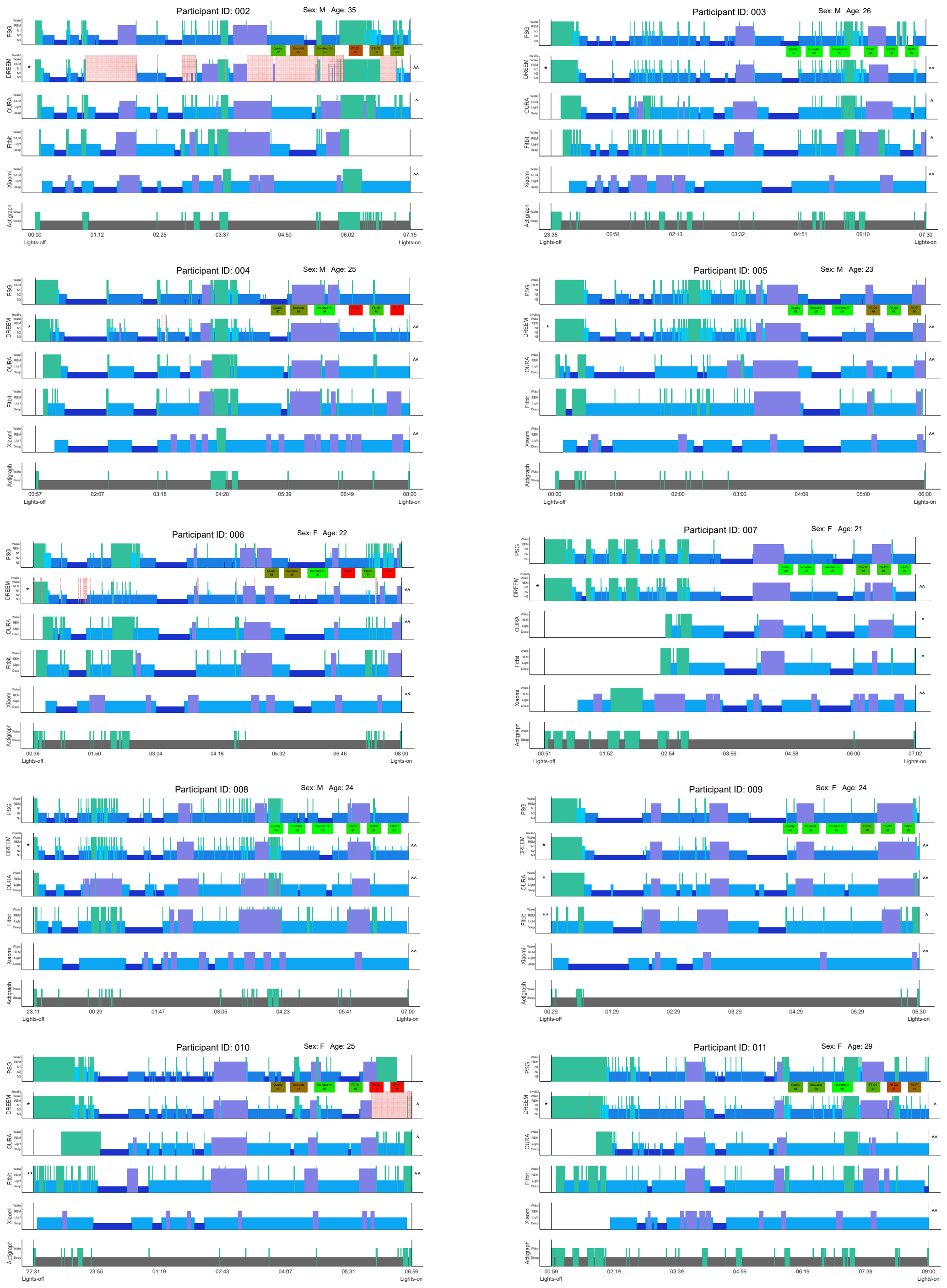

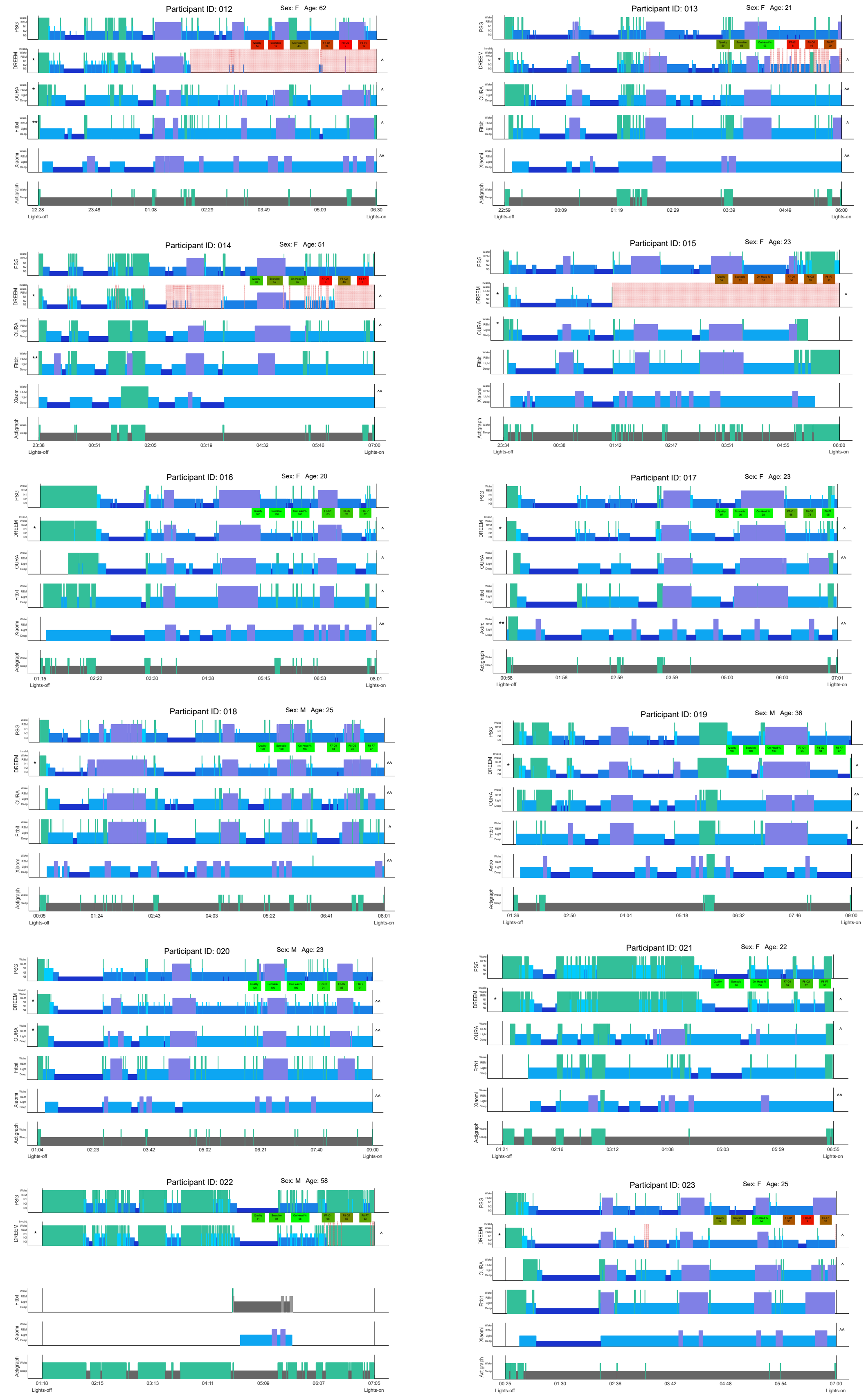
Participant ID: 012
Sex: F Age: 62
Participant ID: 013
Sex: F Age: 21
Participant ID: 014
Sex: F Age: 51
Participant ID: 015
Sex: F Age: 23
Participant ID: 016
Sex: F Age: 20
Participant ID: 017
Sex: F Age: 23
Participant ID: 018
Sex: M Age: 25
Participant ID: 019
Sex: M Age: 36
Participant ID: 020
Sex: M Age: 23
Participant ID: 021
Sex: F Age: 22
Participant ID: 022
Sex: M Age: 58
Participant ID: 023
Sex: F Age: 25

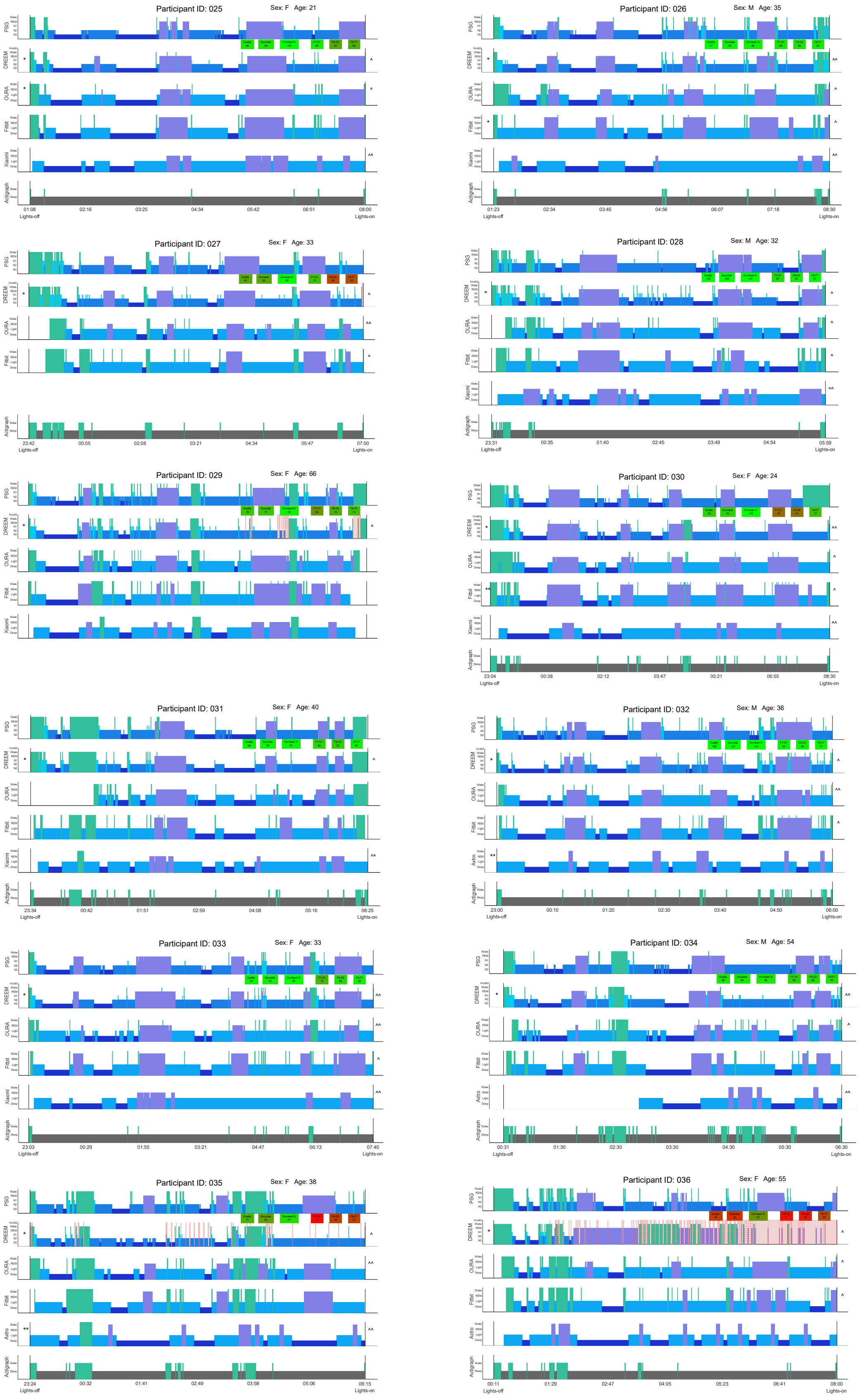
Participant ID: 025
Sex: F Age: 21
Participant ID: 026
Sex: M Age: 35
Participant ID: 027
Sex: F Age: 33
Participant ID: 028
Sex: M Age: 32
Participant ID: 029
Sex: F Age: 66
Participant ID: 030
Sex: F Age: 24
Participant ID: 031
Sex: F Age: 40
Participant ID: 032
Sex: M Age: 36
Participant ID: 033
Sex: F Age: 33
Participant ID: 034
Sex: M Age: 54
Participant ID: 035
Sex: F Age: 38
Participant ID: 036
Sex: F Age: 55

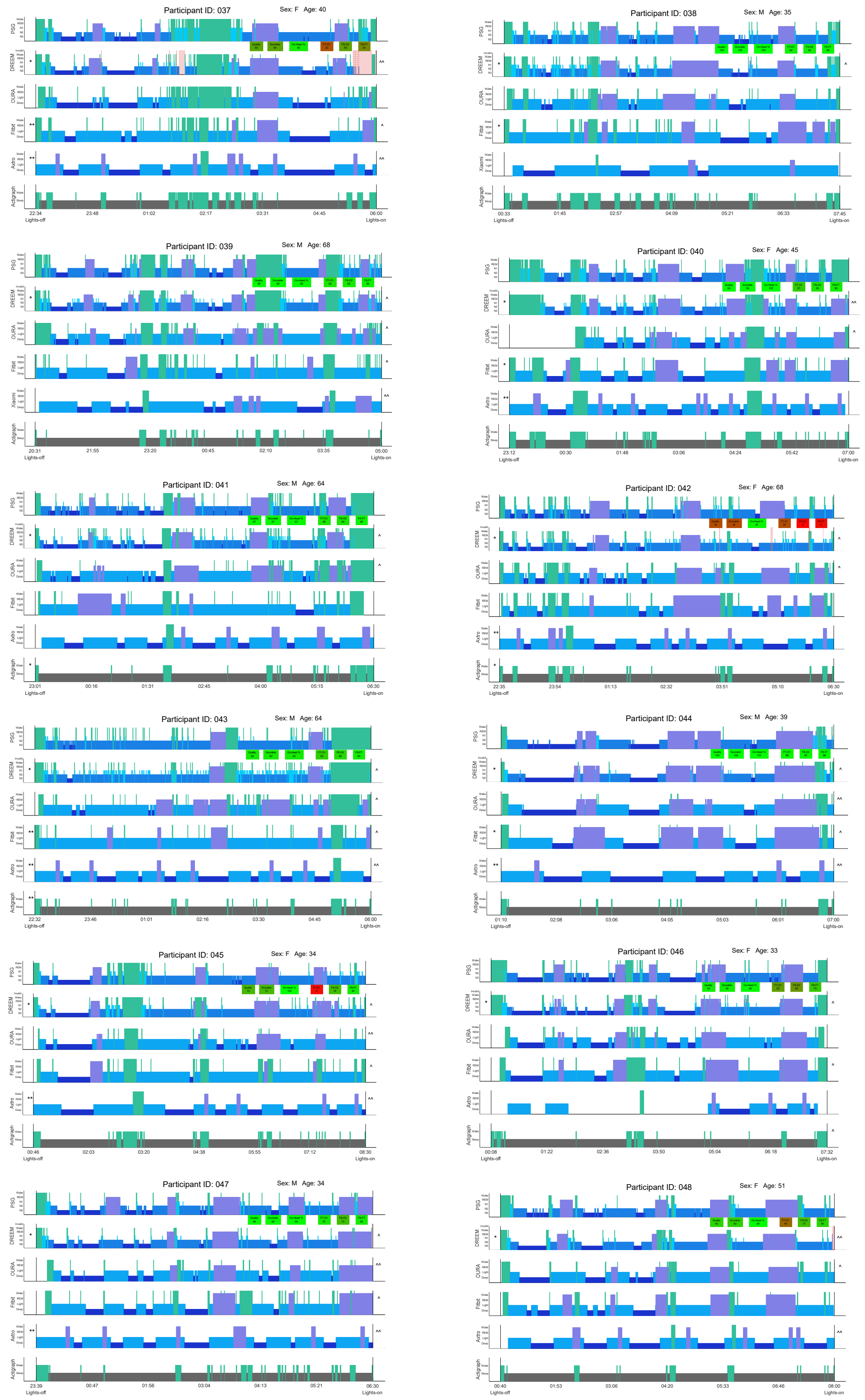

Participant ID: 037 Sex: F Age: 40
Participant ID: 038 Sex: M Age: 35
Participant ID: 039 Sex: M Age: 68
Participant ID: 040 Sex: F Age: 45
Participant ID: 041 Sex: M Age: 64
Participant ID: 042 Sex: F Age: 68
Participant ID: 043 Sex: M Age: 64
Participant ID: 044 Sex: M Age: 39
Participant ID: 045 Sex: F Age: 34
Participant ID: 046 Sex: F Age: 33
Participant ID: 047 Sex: M Age: 34
Participant ID: 048 Sex: F Age: 51
PSG
DREEM
OURA
Fitbit
Axtro
Xiaomi
Actigraph
Lights-off
Lights-on

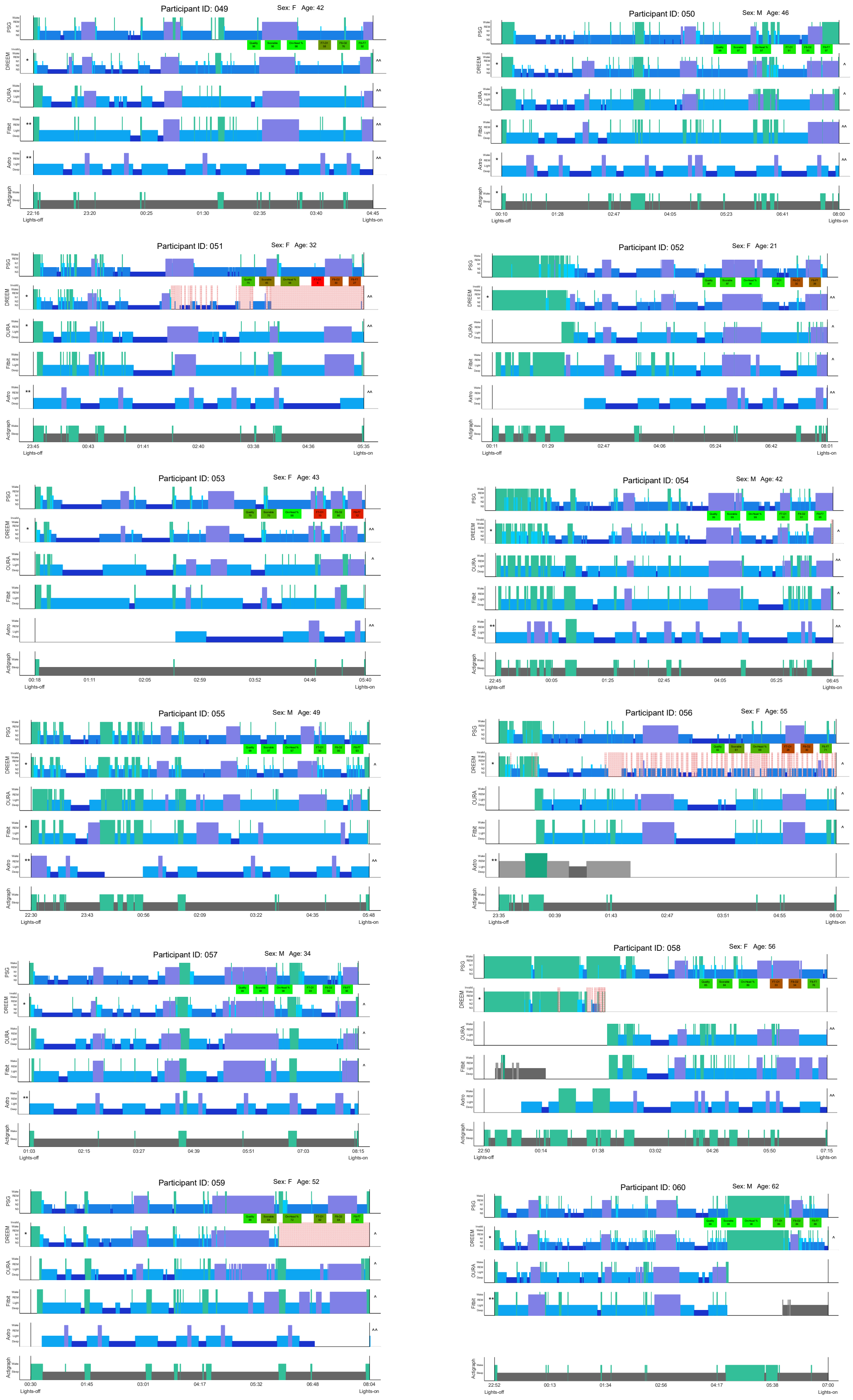
Participant ID: 049
Sex: F Age: 42
Participant ID: 050
Sex: M Age: 46
Participant ID: 051
Sex: F Age: 32
Participant ID: 052
Sex: F Age: 21
Participant ID: 053
Sex: F Age: 43
Participant ID: 054
Sex: M Age: 42
Participant ID: 055
Sex: M Age: 49
Participant ID: 056
Sex: F Age: 55
Participant ID: 057
Sex: M Age: 34
Participant ID: 058
Sex: F Age: 56
Participant ID: 059
Sex: F Age: 52
Participant ID: 060
Sex: M Age: 62

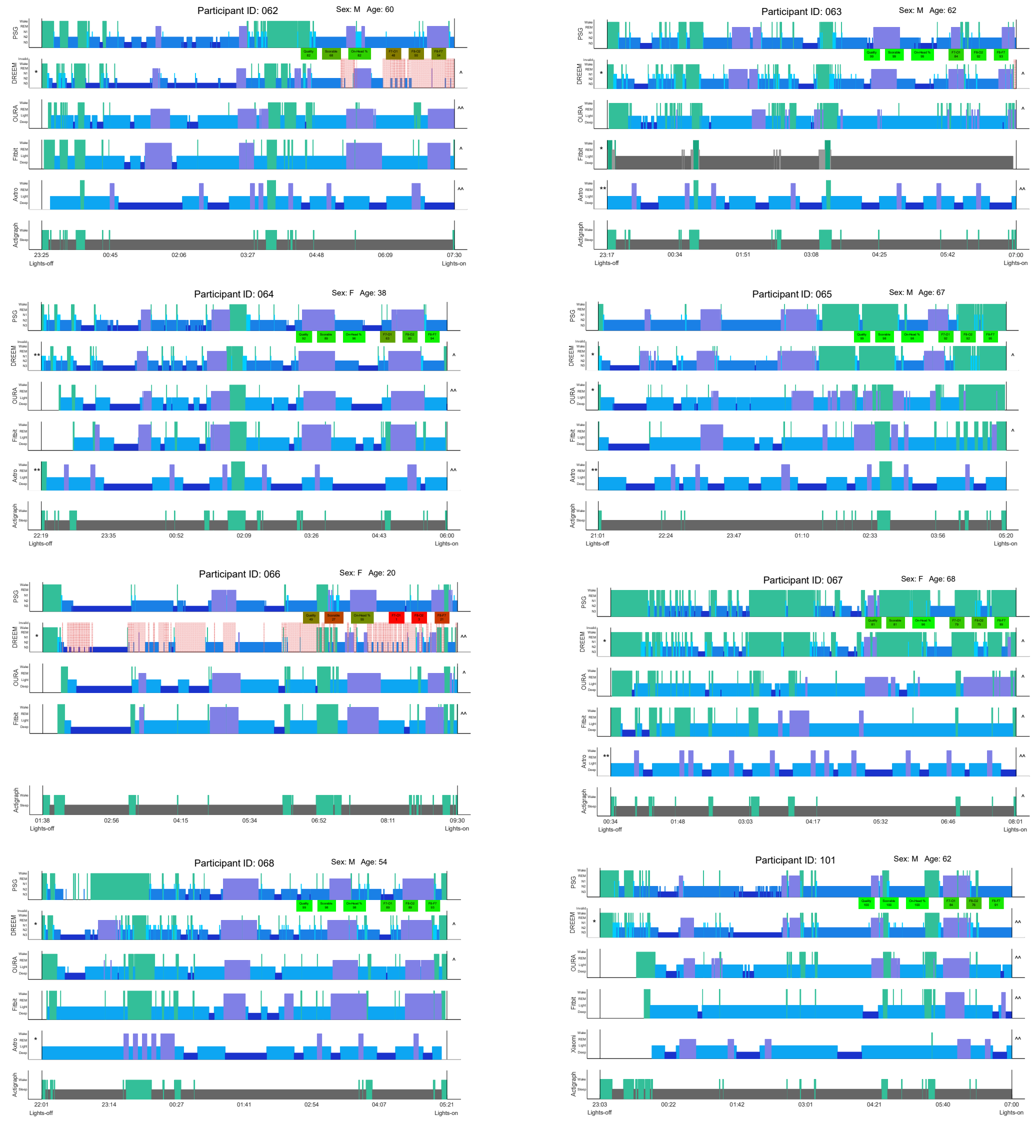


<u>Legend:</u>

| | |
|---|---|
| * | Sleep staging started prior to lights off (only wake staged) |
| ** | Sleep staging started prior to lights off (stages include non-wake stages) |
| ^ | Sleep staging continued post lights on (only wake staged) |
| ^^ | Sleep staging continued post lights on (stages include non-wake stages) |
